# Cancer incidence in first-generation migrants and host populations in seven European registries, including Ukrainian-born migrants in Poland: a population-based study

**DOI:** 10.64898/2026.10.26.26366426

**Authors:** Catharina Johanna Alberts, Damien Georges, Freddie Bray, Alberto Ameijide, Andrea Bordoni, Marià Carulla Aresté, Maria Dolores Chirlaque López, Joanna Didkowska, Jaume Galceran, Koen Van Herck, Armien Lanssens, Enrica Migliore, Laura Ortelli, Lorenzo Richiardi, Antonia Sánchez Gil, Freija Verdoodt, Otto Visser, Urszula Wojciechowska, Andrea Gini, Irene Man, Stefano Rosso, Iacopo Baussano

## Abstract

**Background:** Where people are born shapes their cancer risk, yet European cancer surveillance rarely records migration background. We estimated cancer incidence in first-generation migrants and assessed whether birth-country incidence stratifies their cancer risk.

**Methods:** We harmonised data from six population-based cancer registries in Belgium, the Netherlands, Spain (Murcia, Tarragona), Italy (Turin), and Switzerland (Ticino) for 2003–17, examining breast, colorectal, cervical, stomach, liver, and lung cancer in first-generation migrants and host populations. Countries-of-birth were grouped into quartiles of GLOBOCAN age-standardised incidence by cancer site and sex (*birth-country incidence categories*). We estimated cumulative risks (CR) and migrant-to-host cumulative risk ratios (CRR). Ukrainian-born migrants in Poland were analysed separately for 2022–23.

**Findings:** Migrants’ cancer risk increased with birth-country incidence for cervical, stomach, and liver cancer. For cervical cancer, CRR rose from close to 1 in migrants from low-incidence countries to 4·9 (95% CI 3·5–6·9) in those from high-incidence countries in Turin. Breast and colorectal cancer diagnosis risk was lower than in host populations in every category. Across the six cancers combined, migrants had lower risk in 9 of 11 registry–sex strata, as lower breast, colorectal, and lung cancer risk outweighed higher cervical, stomach, and liver cancer risk. Ukrainian-born migrants in Poland (n=4450) had higher CR at most anatomical sites than the host population.

**Interpretation:** Migrants’ cancer risk relative to host populations depends on country of birth and cancer site. Birth-country incidence is a useful way of grouping migrants by risk. National averages can mask residual risk in subgroups, e.g. migrants born in high-incidence countries had up to five times the cervical cancer risk of host populations with rates near or below the WHO elimination threshold. Routine recording of country of birth can support risk-tailored prevention and cancer service planning for displaced populations.

**Funding:** ZonMw-09150162210124; IARC-WHO; and participating cancer registries.

**RESEARCH IN CONTEXT:** *Evidence before this study:* We searched PubMed, Embase, and Web of Science from inception to Oct 15, 2026, without language restrictions, for population-based studies reporting cancer incidence among first-generation migrants in Europe compared with host populations. We used terms for migration (eg, “migrant”, “immigrant”, “country of birth”), cancer, and incidence. Existing studies specific on migrants in Europe remain sparse and fragmented, with most studies limited to individual countries, migrant groups, or cancer types. They differed in how migrants were grouped, in their reference populations, and in their methods, and often had small numbers per country of birth. A 2010 review reported higher incidence of infection-related cancers among migrants grouped as “non-western”, and similar or lower incidence of breast and colorectal cancer, but heterogeneity prevented quantitative analyses. These differences have been attributed mainly to factors acquired before migration, notably infections, possibly compounded by differences in access to healthcare and preventive services, including lower participation in screening. Comparable estimates across European populations have been lacking. So has evidence on whether birth-country incidence can stratify migrants by cancer risk, and on what registries can capture after recent large-scale displacement.

*Added value of this study:* To our knowledge, this is the first harmonised multi-country analysis to group first-generation migrants by cancer incidence in their country of birth, using data from population-based cancer registries. For cervical, stomach, and liver cancer, cumulative risk among migrants followed the incidence ranking of their countries of birth. For breast and colorectal cancer, it was consistently lower than in host populations, independent of the country of birth. In most settings, the combined burden of the six cancers was lower among migrants. Grouping countries of birth by publicly available national incidence estimates gives a first population-level picture of the cancer profile of migrant populations. A case study of Ukrainian-born migrants in Poland shows what registry data can and cannot analyse and describe after large-scale displacement.

*Implications of all the available evidence:* Routine recording of country of birth in cancer registries can generate evidence for equitable cancer control, supporting planning of prevention, screening and care services for settled and newly arrived migrants, particularly for infection-related cancers. The harmonised protocol and openly available analytic tools developed for this study can be applied by other registries for further research and routine surveillance. Harmonising migration-related data collection across European registries would strengthen surveillance and cancer service planning for displaced populations, and support equity-oriented cancer control policies.

## INTRODUCTION

Europe’s Beating Cancer Plan identifies cancer inequalities as a strategic priority. The European Cancer Inequalities Registry (ECIR), launched in 2022, monitors disparities in cancer prevention, incidence, treatment, and outcomes across multiple socioeconomic dimensions.^1,2^ However, migration background is not currently included, despite migration being an increasingly important determinant of population health. Birth-country information is available in many European cancer registries, either directly or through linkage to civil registration data, although its availability and quality vary considerably between registries.^3^ As the proportion of Europeans living outside their country of birth continues to rise, and population mobility is increasingly shaped by labour migration, conflict, and climate-related displacement, migration background is emerging as an important and under-recognised dimension of cancer inequality.^4,5^

Studies across Europe have consistently shown that first-generation migrants have lower incidence of breast, colorectal, and lung cancer, and higher incidence of cervical, stomach, and liver cancer than host populations.^6,7^ These patterns reflect both exposures acquired before migration (including infectious agents, reproductive factors, diet, tobacco, and alcohol) and post-migration changes both in exposure and in access to prevention, screening, and diagnostic services cancer. Cancer risk may shift towards host-country levels over time. Studies suggest that this shift in risk is gradual and site-specific: risk of breast, colorectal, and lung cancer rises with time since migration, although a risk advantage relative to host populations often persists, whereas the excess risk of infection-related cancers declines only slowly.^6,8–10^ However, existing evidence remains fragmented across countries and migrant groups, limiting comparability and hindering the integration of migration background into European cancer surveillance.

Country-of-origin epidemiology is already being used to inform risk-based screening and prevention strategies for infectious diseases, including hepatitis B and C, HIV, tuberculosis, and parasitic infections.^11^ Whether cancer incidence in migrants’ countries of birth can similarly be used as a valuable indicator of cancer risk after migration has not been systematically assessed across Europe. Establishing whether and when birth-country incidence is informative could support cancer surveillance, prevention, and screening strategies tailored to increasingly diverse populations. The growing emphasis on harmonised European cancer data infrastructures, including the European Health Data Space, EUCanScreen, and the CancerWatch Joint Action, further underscores the need for comparable indicators that capture emerging dimensions of inequality.^12,13^

To address these gaps, we applied a harmonised, semi-distributed analytical approach across seven European population-based cancer registries. We quantified cancer risk among first-generation migrants relative to host populations, assessed whether birth-country incidence can stratify migrants by cancer risk, examined changes across three five-year periods between 2003 and 2017. Furthermore, we investigated cancer incidence among Ukrainian-born migrants in Poland during 2022–23 as a case study of what registry data can and cannot show following large-scale displacement.

## METHODS

In this multi-registry, population-based analysis, we compared cancer incidence among first-generation migrants and host populations. The study was developed within the Cancer RADAR initiative, which has established a standardised analytical workflow for migration-stratified analyses of cancer incidence across European population-based cancer registries (PBCRs).^3^

Data over the period 2013-17 were contributed by six PBCRs, covering Belgium, the Netherlands, Spain (Murcia and Tarragona), Italy (Turin), and Switzerland (Ticino), and by a seventh registry, in Poland, for Ukrainian-born migrants over 2022–23. Data were grouped into three 5-year periods corresponding to successive volumes of Cancer Incidence in Five Continents: 2003–07 (CI5-X), 2008–12 (CI5-XI), and 2013–17 (CI5-XII).^14^ Data from the Polish registry covered incident cases diagnosed in 2022–23 and were analysed separately. We analysed breast cancer (ICD-10 C50), colorectal cancer (C18–C20), cervical cancer (C53), stomach cancer (C16), liver cancer (C22), lung cancer (C33–C34; C34 only in Belgium), and all cancer sites combined (C00–C97, excluding C44).

The PBCRs identified first-generation migrants either directly from registry records or, in Belgium, through linkage with census data from Statistics Belgium (Statbel; see Supplementary Appendix A.2).^15^ The type and availability of data are summarised in **Supplementary Table 1A**. Data definitions and quality checks followed the IARC Cancer Incidence in Five Continents series.^14^ The data-quality indicators collected are described in the **Supplementary Appendix A.1.**

First-generation migrants were defined as residents of the registry catchment area whose country of birth differed from the country in which the registry was located, consistent with the UN operationalisation of international migrants on the basis of foreign-born status.^16^ The term is equivalent to “foreign-born” and refers solely to country of birth, irrespective of nationality, citizenship, or duration of residence, which can change over time. The host population was defined as individuals born in the registry country. Second-generation migrants (individuals born in the host country with one or both parents born abroad) could not be identified separately and were therefore included in the host population. Country of birth was coded using ISO 3166 codes.^17^ Individuals with an unknown or unrecorded country of birth were recorded separately for data-quality reporting but were excluded from the analyses.

Countries of birth were grouped into quartiles of national age-standardised incidence rates (ASRs), obtained from GLOBOCAN 2022, hereafter referred to as birth-country incidence categories (low, intermediate-low, intermediate-high, and high).^18^ Quartile boundaries were set separately for each cancer site and sex, so a given country may fall into different categories across sites and/or sexes (**Supplementary Table 1B**). These categories describe the background cancer burden in the country of birth and are not attributes of individual migrants. The countries of birth represented in each category, by registry and cancer site, are described in **Supplementary Table 1C.**

### Belgium-specific approach

In Belgium, first-generation migrant status was identified through linkage with the 2001 national census for adult residents 18 years or older and covering 95% of the population.^15^ Because this closed cohort aged during follow-up, Belgian cumulative risks (CRs) were calculated over progressively older, truncated age ranges and therefore slightly underestimate full-range risk. Belgium contributed data from 2004 onwards and only to the six site-specific cancer analyses. ASRs were not calculated for Belgium. Full details are provided in **Supplementary Appendix A.2**.

### Risk and effect measures

All analyses were run locally at each registry using the cancerradarr R package developed for the Cancer RADAR initiative.^19^ Cumulative risk (CR) from age 20 to 74 years was the primary outcome measure, calculated from age-specific incidence rates in 5-year age groups. Shared registry data start at age 20, and for Belgium the lower age limit was defined by the census cohort (30 years in 2013–17). CR does not depend on an external standard population and remains interpretable over a truncated age range, allowing Belgian estimates to be presented alongside those of the other registries. ASRs are reported in the supplementary material because they remain the standard measure for international comparison and are the scale on which the WHO cervical cancer elimination threshold is defined. Birth-country CRs and ASRs were obtained from GLOBOCAN 2022.^18^ CRs were estimated for the host population, for each birth-country incidence category and, in supplementary analyses, for each Human Development Index (HDI) category (low, medium, high, and very high)^20^ and UN region as defined in GLOBOCAN^18^.

CR ratios (CRRs) and CR differences (CRDs) comparing first-generation migrants with the host population were calculated with corresponding 95% confidence interval (CI). For absolute-difference analyses, CRDs were pooled across all first-generation migrants relative to the host population within each registry, by sex and cancer site. All main analyses (CRs, CRRs, and CRDs) used the most recent period common to all registries (2013–17). Time-trend analyses used all available periods: three for most registries and four for Belgium (including 2018–22). Ticino was not included in these analyses because population-at-risk data were available for only one period (2013–17). Trends were quantified as the change in the CRR between periods, with 2013–17 as reference. Because the Belgian age window shifts upwards between periods, Belgian trend estimates compare slightly different age ranges.

### Software, data protection, and ethics

All analyses were performed using R version 4.6.0. The Cancer RADAR analytical pipeline (cancerradarr R package, version 3.0.0) generated standardised aggregated outputs locally within each registry.^19^ Only aggregated outputs (risk indicators and their 95% CIs) were shared by registries through the IARC-hosted REDCap platform. To protect confidentiality, strata with fewer than five cancer cases or fewer than 100 person-years were combined with adjacent age bands to protect confidentiality, separately for each registry, sex, cancer site, and migrant group. Estimates based on fewer than five cases after aggregation were not provided in the output file. The study was approved by the IARC Ethics Committee (approval number IEC 23-38). Each registry contributed aggregated data in accordance with their applicable national legislations.

## RESULTS

The total number of cancer cases during 2013–17 ranged from 11 510 in Ticino to 497 324 in the Netherlands (**Supplementary Table 2**); Belgium did not provide an all-cancers count. In four registry regions, migrants comprised 10·9–17·9% of the source population but accounted for only 3·7–8·0% of all cancer cases, a difference that largely reflects the younger age structure of migrant populations and their lower risk. Ticino was an exception: there the migrant population had an age structure similar to that of the host population (48% aged under 50 in both), and migrants accounted for 38–40% of cases against 35–36% of the population.

For cervical, stomach, and liver cancer, pooled CR among migrants followed the same order as the birth-country incidence categories (**figure 1**). Risk was higher among migrants born in higher-incidence countries, with the highest CRRs in the high-incidence category (**figure 2**). In other words, the ordering of countries by their background incidence was largely preserved after migration. The gradient was steepest for cervical cancer in females, for which pooled CRRs were close to 1 among migrants born in low-incidence countries and reached 4·88 (95% CI 3·48–6·85) among those born in high-incidence countries in Turin and 6·86 (3·58–13·12) in Murcia, although the latter estimate was based on fewer cases (**figure 2**). There were two exceptions: in the Netherlands, risk among migrants born in high-incidence countries was close to that of the host population, and in Belgium no gradient was observed (see footnote **figure 2** and **Discussion**). Cervical cancer ASRs in the host population were below the WHO elimination threshold of 4 per 100 000 person-years in Turin (3·9) and Tarragona (3·3), and above but approaching it in the other registries (**Supplementary Figure 1**). Similar gradients were seen for stomach cancer in both sexes (up to CRR 5·92 [95% CI 3·97–8·84] among males born in high-incidence countries in Turin) and liver cancer in males (up to 3·17 [2·66–3·78] in the Netherlands).

**Figure 1:**
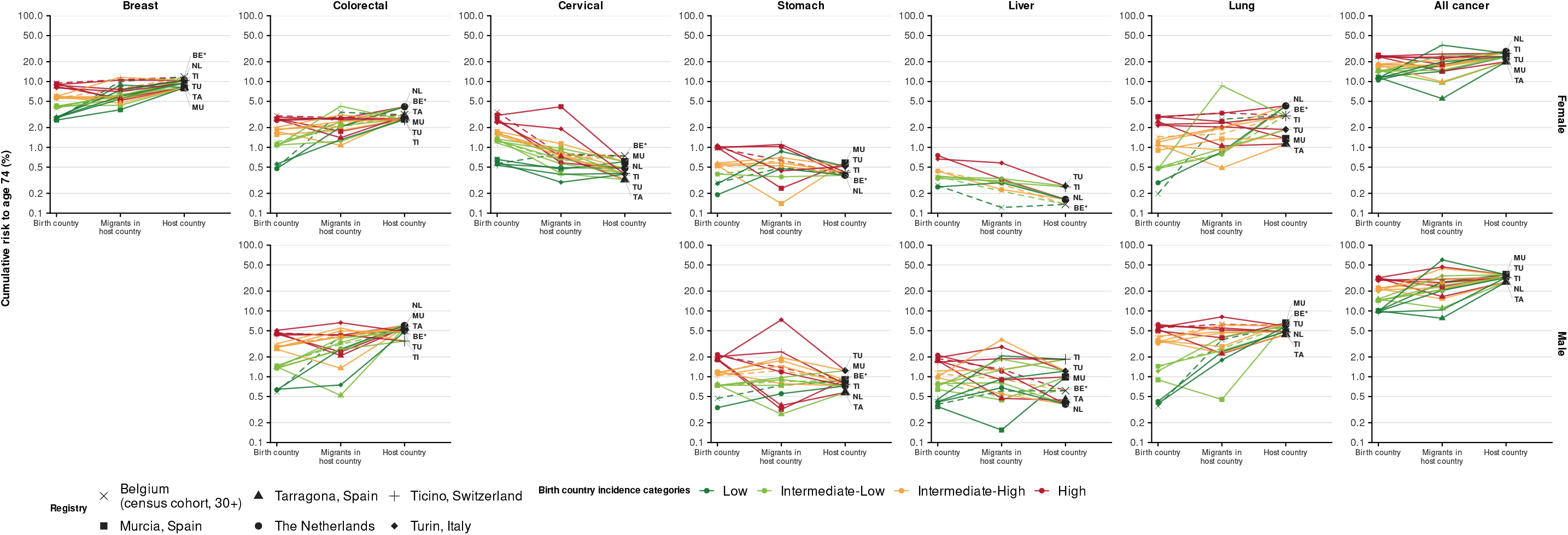
Cumulative risk (20–74) of first-generation migrants in relation to birth-country and host-country risk, by birth-country incidence category, cancer site, and sex, in six European population-based cancer registries, 2013–17. CR = cumulative risk from age 20 to 74 years (%), log scale. Each panel shows one cancer site; within each registry, four lines correspond to the four birth-country incidence categories, defined separately for each cancer site and sex from GLOBOCAN 2022 age-standardised incidence rates (**Supplementary Table 1B**), and each symbol marks one registry. The three categories on the X axis are **(1) birth-country** = birth country CR, a person-year-weighted average of the GLOBOCAN 2022 cumulative risks of the countries of birth in that category, over the same age range; **(2) migrants in host country** = the observed CR among first-generation migrants in the registry; and **(3) host country** = the observed CR in the host population. All panels share one Y axis range. Strata with fewer than 5 cases are not shown. **Note:** Belgium is represented with a dashed line. In Belgium, first-generation migrant status was obtained through linkage with the 2001 national census, restricted to individuals aged ≥18 years at the census baseline (1 October 2001). Because this closed cohort was followed forward, the observable age range is incomplete and shifts towards older ages over time. Belgian CRs for 2013–17 are therefore age-truncated to ages 30–74 years. They remain comparable with those of the other registries but underestimate full-range risk.

**Figure 2:**
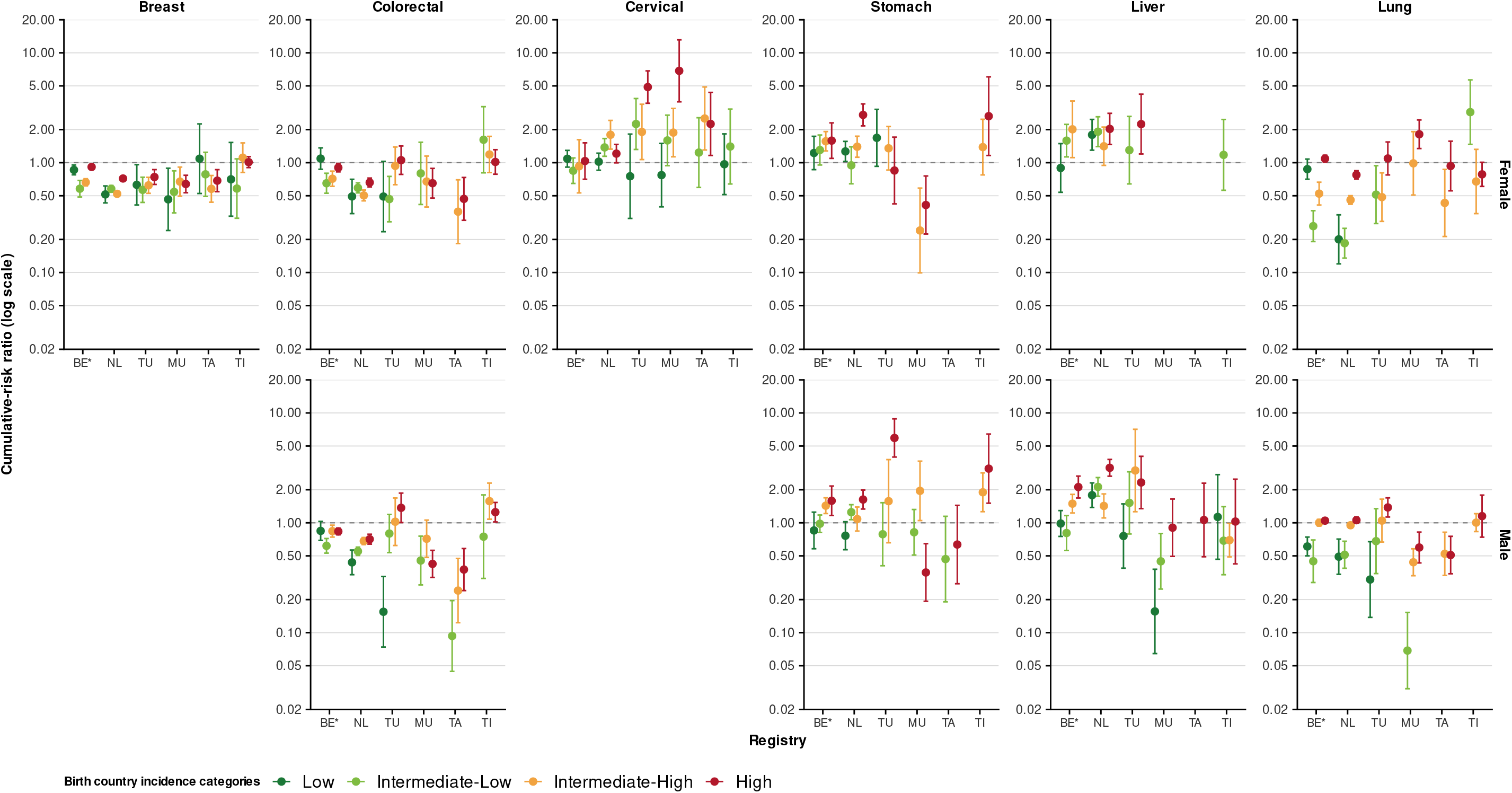
Cumulative risk ratios (20–74) comparing first-generation migrants with the host population, by birth-country incidence categories, cancer site, and sex, in six European population-based cancer registries, 2013–17. CRR = cumulative risk ratio (migrants vs host), based on cumulative risk from age 20 to 74 years. BE = Belgium; NL = the Netherlands; TU = Turin, Italy; MU = Murcia, Spain; TA = Tarragona, Spain; TI = Ticino, Switzerland. Within each panel, registries are on the X axis and the four birth-country incidence categories are shown as coloured dots with 95% CI bars; the Y axis is on a log scale, and the dashed horizontal line marks a CRR of 1. Strata with fewer than 5 cases are not shown. Breast and cervical cancer are shown for females only. **Interpretation:** For cervical, stomach, and liver cancer, CRRs were generally highest among migrants born in high-incidence countries. For breast and colorectal cancer, CRRs were below 1 in most category. For lung cancer, CRRs were mostly below 1 among migrants born in countries below the global median incidence, whereas among those born in countries above it, they ranged from 0·43 to 1·81 and were close to 1 in half of the categories. In a few categories, countries of birth with very different risks were pooled, so the category estimate lay between them. The clearest examples was **cervical cancer** in the high-incidence category, where the largest groups were close to the host level: Surinamese-born (CRR 1·41, 95% CI 1·09–1·83) and Indonesian-born women (1·25, 0·79–1·97) in the Netherlands, and Congolese-born women in Belgium (0·74, 0·44–1·24). By contrast, Romanian-born women had a four- to sevenfold excess in all three registries where they were reported (Turin 6·75, 4·61–9·87; Netherlands 4·27, 2·17–8·42; Belgium 4·09, 2·22–7·53). Another example was for **liver cancer** in Dutch men, where the category estimate of 3·17 (2·66–3·78) combined Indonesian-born men (2·06, 1·38–3·08) with Vietnamese-born (11·97, 7·43–19·29), Egyptian-born (8·33,5·15–13·47) and Chinese-born (7·08, 4·83–10·39).

Migrants had lower risk of breast and colorectal cancer than the host population independent of their country of birth (**figure 1**). For breast cancer, 21 of 24 CRRs across registries and categories were below 1 (**figure 2**). For colorectal cancer, CRRs were below 1 in 15 of 20 categories in females and 17 of 21 in males. No CRR for breast cancer was significantly above 1, and for colorectal cancer only three were, all among males (Turin and Ticino). (**figure 2**).

For lung cancer, the pattern depended on birth-country incidence category. Migrants from countries below the global median ASR (the low and intermediate-low categories) had lower pooled CR than the host population, with CRRs significantly below 1 in 10 of 13 categories. The exceptions were females in the lowest category in Belgium (CRR 0·87, 95% CI 0·71–1·08) and two categories based on eight cases each in Turin and Ticino. Among migrants from countries above the global median, the CR was similar to that of the host population in 12 of 24 categories (**figure 2**).

**Supplementary Figures 2A–2B** present the CRRs grouped by HDI category and UN region: cervical cancer CRRs were highest among migrants from very-high-HDI countries in Belgium, Tarragona, and Turin.

In absolute terms, the lower risk of breast, colorectal, and lung cancer among migrants compared with to the host population outweighed their higher absolute risk of cervical, stomach, and liver cancer: the net CRD over the six cancers was negative in 9 of the 11 registry and sex strata, and was positive only among males in Turin (+3·9%) and Ticino (+1·1%) (**figure 3**). Relative and absolute differences diverged: the largest CRRs were observed for cervical cancer, but CRD did not exceed 0·8% (Turin), whereas breast and colorectal cancer had lower CRRs but larger absolute differences, of up to −3·9% (breast, females, the Netherlands) and −4·1% (colorectal, males, Tarragona).

**Figure 3:**
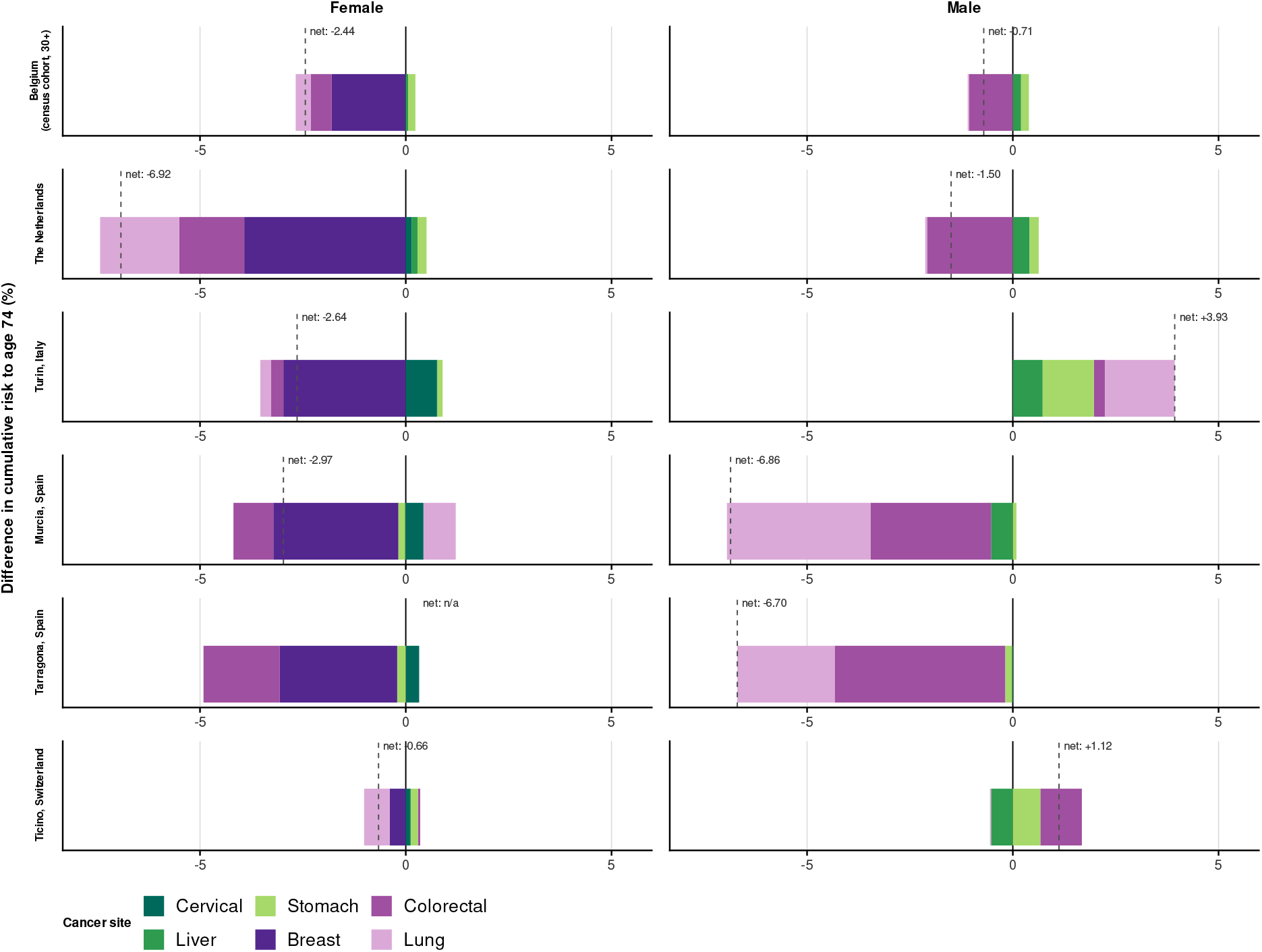
Pooled cumulative risk differences (CRDs) (20–74) between first-generation migrants and host populations, by registry, sex, and cancer site, in six European population-based cancer registries, 2013–17. CRD = cumulative risk difference (20–74, in percentage points). Each bar is one registry and sex; coloured segments are the site-specific CRDs, stacked with negative values (lower risk among migrants) to the left of zero and positive values to the right. The dashed vertical line and label give the net CRD, the sum of the six site-specific CRDs in females and of four in males. A net is shown only when every cancer site has at least 5 cases; it reads “net: n/a” for females in Tarragona, where the liver cancer had was suppressed. **Interpretation:** The lower absolute risk of breast, colorectal, and lung cancer among migrants generally outweighed their higher risk of cervical, stomach, and liver cancer, so net CRDs were negative in 9 of the 11 strata with an estimable net.

Few changes were observed in CRRs over the time periods analysed, with no clear sign of convergence towards host-population risk (**Supplementary Figure 3**, **Supplementary Table 3**). However, confidence intervals were wide in the smaller registries, and moderate changes cannot be excluded. In Murcia and Tarragona, the significant changes moved CRRs for some cancer sites (e.g., liver and stomach in men) that had been below 1 closer to 1, largely reflecting increasing risk among migrants. In the Netherlands, significant changes were small and mainly reflected changes in host-population risk, with CRRs for breast and colorectal cancer moving further below 1 and those for stomach cancer increasing.

Data-quality indicators by registry, sex, cancer site, and migration status are presented in **Supplementary Table 4**. They showed good to high data quality across all cancer registries and type of cancer types, without significant differences between migrant and host populations. Missingness of birth-country information was generally low, except in Tarragona, where it reached up to a third of cases for some sites (stomach, cervical, liver, and lung cancer).

Among Ukrainian-born migrants in Poland, 4450 incident cancer cases were recorded during 2022–23, of which 67% (n=2995) were in females. In females, CR was broadly similar to that in Ukraine for colorectal, cervical, stomach, and lung cancer, but higher for breast cancer (**figure 4.A**). In males, CR was higher than both in Ukraine and in the host Polish population at all four sites (**figure 4.A**). Relative to the host Polish population, CR was significantly higher in 8 out of 10 cancer site and sex specific combinations (**figure 4.B**). The age and sex structure of the Ukrainian-born population differed substantially from those of the Ukrainian and host Polish populations with a younger population and an overrepresentation of females compared to both the host Polish population and Ukrainian general population (**Supplementary Figure 4**).

**Figure 4:**
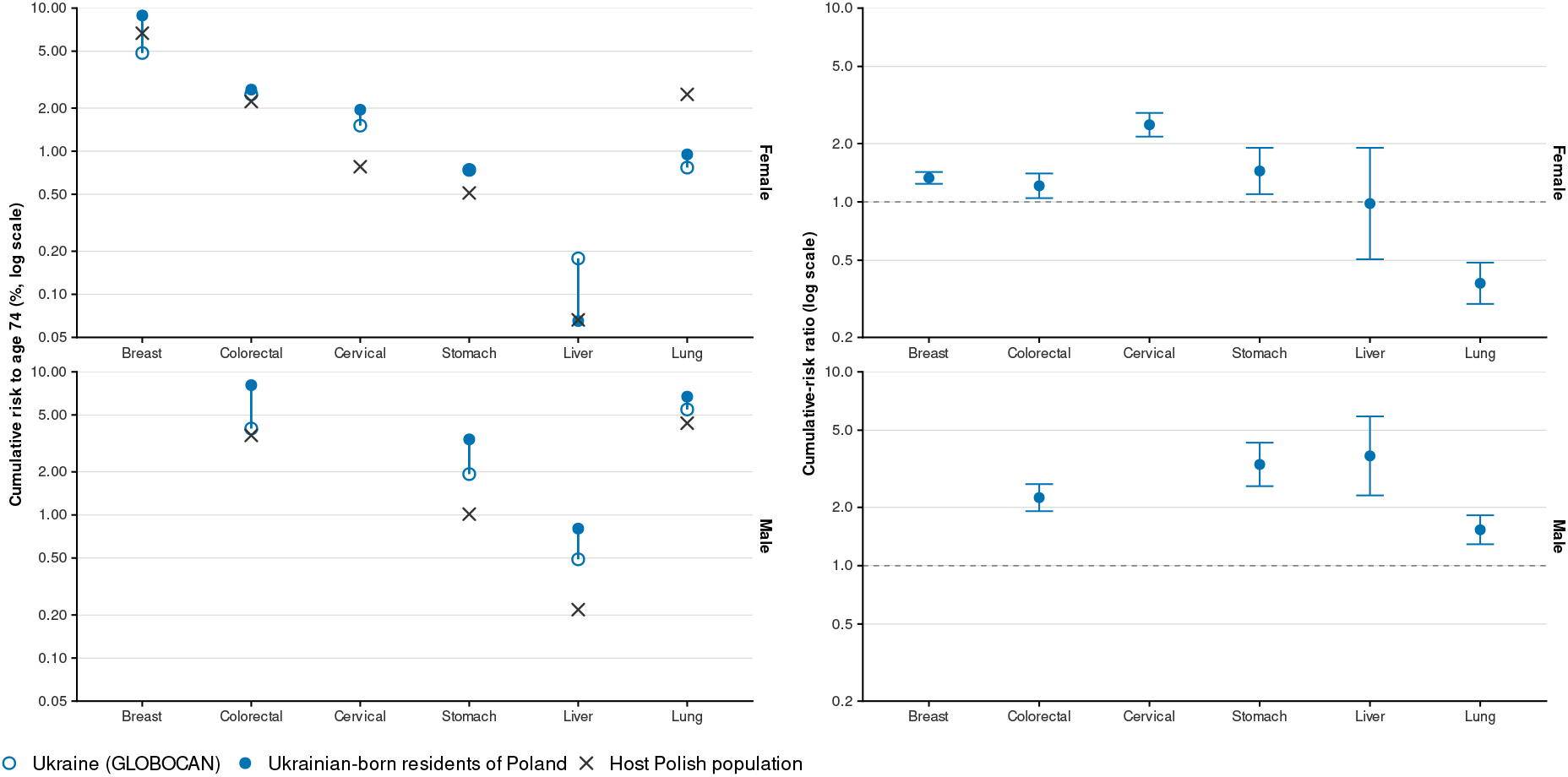
Cumulative risk among Ukrainian-born residents of Poland in relation to cumulative risk in Ukraine and in the host Polish population (left) and cumulative risk ratios comparing Ukrainian-born residents of Poland with the host Polish population (right), by cancer site and sex, Polish National Cancer Registry, 2022–23. CR = cumulative risk from age 20 to 74 years (%), log scale; both panels share one Y axis range. For each cancer site, the open circle is the CR in Ukraine (GLOBOCAN 2022 estimate over the same age range), the filled circle the observed CR among Ukrainian-born residents of Poland, and the cross the observed CR in the host Polish population. The line joins Ukraine to the Ukrainian-born residents of Poland. CRR = cumulative risk ratio, based on cumulative risk from age 20 to 74 years. Points show the CRR and bars its 95% CI; the Y axis is on a log scale, and the dashed line marks a CRR of 1 (equal risk). Estimates are based on 4450 cancer cases among Ukrainian-born residents (2995 in females, 1455 in males); every site had at least 5 cases. Breast and cervical cancer are shown for females only. **Interpretation:** In females, cumulative risk among Ukrainian-born residents of Poland was close to that in Ukraine for colorectal (2·69% vs 2·50%), cervical (1·95% vs 1·51%), stomach (0·74% vs 0·74%), and lung cancer (0·95% vs 0·77%), and above both the Ukrainian and the native Polish levels for breast cancer (8·88% vs 4·86% and 6·68%). In males, it was above both at all four sites, most markedly for colorectal (8·07% vs 4·02% and 3·60%) and stomach cancer (3·37% vs 1·93% and 1·01%). Cumulative risk among Ukrainian-born residents of Poland was higher than in the host Polish population for most cancer sites. In males, risk was higher at all four sites, most markedly for liver (CRR 3·69, 95% CI 2·30–5·89) and stomach cancer (3·33, 2·57–4·31). In females, the largest excess was for cervical cancer (2·50, 2·17–2·88), followed by stomach, breast and colorectal cancer. The exceptions were lung cancer in females, where risk was lower (0·38, 0·30–0·49), and liver cancer in females, where it did not differ (0·98, 0·51–1·90).

## DISCUSSION

This study provides the first empirical results from the Cancer RADAR initiative^3^, with harmonised estimates of cancer incidence among first-generation migrants across European population-based cancer registries. There are four key findings. First, CR among migrants largely followed the incidence ranking of their countries of birth for cervical, stomach, and liver cancer. For breast and colorectal cancer, a weaker ranking was present, but CR was lower than in host populations in every category, and for lung cancer the ranking was weaker and inconsistent. Second, the combined burden of the six cancers studied was lower among migrants than among host populations in most registry and sex strata. Third, CRRs were largely stable over time, with modest changes in some registries. Lastly, among Ukrainian-born migrants in Poland, CR was higher than in the host population for most cancer sites and both sexes.

The different patterns across cancer types suggest that the timing and nature of the relevant exposures vary by site. For cervical, stomach, and liver cancer, risk among migrants followed the incidence ranking of their countries of birth, indicating that factors acquired before migration, notably human papillomavirus, *Helicobacter pylori*, and hepatitis B and C infection, continue to influence risk after arrival.^6,10,21^ These may be compounded by differences in access to healthcare and preventive services.^22,23^ For lung cancer, risk remained low among migrants from low-incidence categories but was less consistent in high-incidence categories, probably reflecting the stage of the smoking epidemic in the birth country.^24^ For breast and colorectal cancer, a weaker ranking by birth-country incidence was present, but risk was lower than in host populations in every category. This may reflect unmeasured factors as changes in reproductive patterns and lifestyle, which outweigh differences in birth-country incidence levels.^7^ However, lower screening participation may also reduce the detection of breast cancer, and to a lesser extent colorectal cancer, among migrants.

Two exceptions illustrate how category composition can shape these estimates. In the Netherlands, the high-incidence cervical cancer category is dominated by long-established Surinamese-born and Indonesian-born populations, whose risk has largely converged towards that of the host population (CRRs 1·41 and 1·25), despite a fourfold excess among Romanian-born women in the same category (4·27). In Belgium, the gradient was compressed from both ends: women born in neighbouring low-incidence countries had elevated risk (CRRs 1·31–1·40), while the high-incidence category was dominated by the long-established Congolese-born population, whose risk did not differ significantly from that of the host population (0·74, 95% CI 0·44–1·24), rather than by Romanian-born women in the same category (4·09), leaving no gradient to observe.

Our findings are consistent with recent multi-country studies from the Nordic countries that reported similar patterns as observed within Cancer RADAR of incidence among migrant populations, although those analyses grouped migrants by broad geographic regions of origin rather than country of birth.^8,21^ Our analysis extends these observations to a different set of registries, includes both sexes, and reports absolute alongside relative measures of burden.

These findings have important implications for cancer prevention and planning. The gradient in risk across birth-country incidence categories for cervical, stomach, and liver cancer supports equity-focused interventions for populations born in high-incidence settings: greater participation of migrant groups in HPV-based cervical screening, more targeted *Helicobacter pylori* screen-and-treat strategies, and screening for hepatitis B and C with linkage to care. Although cervical cancer rates in the host populations of some registries were below or close to the WHO elimination threshold of 4 per 100 000 person-years, CR among migrants born in high-incidence countries was several times higher. Thus, national incidence approaching the threshold may obscure subgroups well above it, and monitoring by migration background is needed to ensure that elimination is achieved equitably rather than only on average. For breast and colorectal cancer, equitable participation in organised screening programmes remains essential as migrant populations age, even though their risk is currently lower than that of host populations.

Reporting both relative and absolute measures is important. Risk ratios identify groups at higher or lower risk and inform discussions of cause, whereas risk differences quantify disease burden and are more directly relevant to national service planning. The predominantly negative net CRDs challenge the idea that migrants consistently experience a higher cancer burden and support a cancer-specific approach to planning and prevention. CR also removes confounding by age structure, which matters here because migrant populations are considerably younger than host populations.

The Ukrainian case study shows what registry data can and cannot establish following displacement, a setting in which a lack of quantitative information is a recognised challenge for humanitarian cancer programmes.^25,26^ There are several possible explanations for the varying risk relative to the host Polish population: (i) differences in registration coverage and diagnostic capacity between Ukraine and Poland, (ii) care-seeking migration among people already in treatment, (iii) genuine differences in pre-migration exposure, and (iv) challenges in capturing denominator data of recently displaced populations. Distinguishing them requires data on age at migration, date of arrival, or duration of residence. The predominantly female population is consistent with displacement after February 2022, since men aged 18 to 60 years were largely unable to leave Ukraine, whereas the men present are more likely to be pre-war labour migrants, estimates for males should therefore not be read as reflecting risk on arrival.

Strengths include the semi-distributed approach, in which registries analysed individual-level data locally and shared aggregated results, ensuring comparable analyses while respecting data protection, and the reporting of data-quality indicators alongside the results.

Several limitations should be considered. Migrants were classified only by country of birth, which does not capture variation within their countries of birth. Incidence estimates for countries of birth may be uncertain where registration is less complete, which affects comparisons with birth-country risk reported here. Strengthening population-based cancer registration globally^27^ is therefore critical for monitoring risk in increasingly mobile populations. Aggregated data lacked information on socioeconomic position, duration of residence, age at migration, and screening participation, all of which may influence risk. Country of birth was missing for up to a third of cases for some sites in Tarragona. In addition, second-generation migrants could not be identified separately and were included in the host population, which is likely to have attenuated the observed differences between migrants and host populations.^9^

In conclusion, cancer risk among first-generation migrants in Europe largely reflected incidence in their countries of birth for cervical, stomach, and liver cancer and was lower than in host populations for breast and colorectal cancer. Recently displaced populations, such as Ukrainian-born migrants in Poland, may deviate from this pattern. Harmonised, migration-stratified analysis of registry data is feasible and can inform more equitable cancer control across Europe.

## CONTRIBUTORS

CJA, IB, DG, SR, and FB conceived the study. All authors contributed to the development of the Cancer RADAR protocol and the standardised data collection tool. DG, CJA, SR, and IB developed the cancerradarr R package. AA, AB, MCA, MDCL, JD, JG, KVH, AL, EM, LO, LR, ASG, SR, FV, OV, and UW collected and cleaned the data at registry level, ran the cancerradarr package, and shared the aggregated outputs with the Cancer RADAR team. DG and CJA accessed and verified the underlying data reported in the manuscript. CJA wrote the first draft of the manuscript. AG and IM contributed to the interpretation of the results. All authors contributed to the interpretation of the data, critically revised the manuscript, and approved the final version. Registry-based authors had full access to the data from their own registry. DG and CJA had full access to all the pooled aggregated data in the study. All authors had final responsibility for the decision to submit for publication.

## DATA SHARING

Requests for data access can be submitted to IB and will then be forwarded to each individual registry. The purpose of the research will be carefully evaluated, and a detailed, well-founded justification will be required to prevent any misuse of the data.

## ACKNOWLEDGMENTS

The Cancer RADAR project was funded by a VENI fellowship from the Netherlands Organisation for Health Research and Development (ZonMw; grant number 09150162210124), which covered personnel costs for CJA. Participating cancer registries contributed local resources for data retrieval and linkage. The International Agency for Research on Cancer (IARC/WHO) provided additional personnel, computing resources, and supported data collection. AG is supported by the HPV FASTER Implement project European Health and Digital Executive Agency (HADEA), grant number 101155975. We thank Maarten F Schim van der Loeff, Public Health Service of Amsterdam, the Netherlands, for critically reading the manuscript and for valuable feedback on the project.

Where authors are identified as personnel of the International Agency for Research on Cancer/WHO, the authors alone are responsible for the views expressed in this article, and they do not necessarily represent the decisions, policy, or views of the International Agency for Research on Cancer/WHO.

