## Supplementary Materials for "Cancer incidence in first-generation migrants and host populations in seven European registries, including Ukrainian-born migrants in Poland: a population-based study"


**AUTHOR INFORMATION:**

Catharina Johanna Alberts, Damien Georges, Freddie Bray, Alberto Ameijide, Andrea Bordoni, Marià Carulla Aresté, Maria Dolores Chirlaque López, Joanna Didkowska, Jaume Galceran, Koen Van Herck, Armien Lanssens, Enrica Migliore, Laura Ortelli, Lorenzo Richiardi, Antonia Sánchez Gil, Freija Verdoodt, Otto Visser, Urszula Wojciechowska, Andrea Gini, Irene Man, Stefano Rosso, Iacopo Baussano

- Department of Epidemiology and Data Science, Amsterdam UMC, Amsterdam, Netherlands (C.J. Alberts PhD)
- Public Health Service Amsterdam, Amsterdam, Netherlands (C.J. Alberts PhD)
- Early Detection, Prevention and Infections Branch, International Agency for Research on Cancer, Lyon, France (C.J. Alberts PhD, D. Georges MSc, A. Gini PhD, I. Man PhD, I. Baussano MD PhD)
- Cancer Surveillance Branch, International Agency for Research on Cancer, Lyon, France (F. Bray PhD)
- Tarragona Cancer Registry, Cancer Epidemiology and Prevention Service, Hospital Universitari Sant Joan de Reus, Institut de Recerca Biomèdica Catalunya Sud (IRBCatSud), Reus, Spain (A. Ameijide MSc, M. Carulla Aresté MSc, J. Galceran PhD)
- Ticino Cancer Registry, Public Health Division, Health Department, Locarno, Switzerland (A Bordoni PhD, L Ortelli MSc)
- Murcia Cancer Registry, Department of Epidemiology, Murcia Regional Health Council, IMIB-Arrixaca, Murcia, Spain (M.D. Chirlaque López MD PhD, A. Sánchez Gil MD)
- CIBER of Epidemiology and Public Health (CIBERESP), Madrid, Spain (M.D. Chirlaque López MD PhD)
- Polish National Cancer Registry, Maria Sklodowska-Curie National Research Institute of Oncology, Warsaw, Poland (J. Didkowska PhD, U. Wojciechowska PhD)
- Belgian Cancer Registry, Brussels, Belgium (K. van Herck MD PhD, A. Lanssens PhD, F. Verdoodt PhD)
- Piedmont Cancer Registry, University Hospital Città della Salute e della Scienza di Torino and CPO Piemonte, Turin, Italy (E. Migliore PhD, Prof L. Richiardi PhD, S. Rosso PhD)
- Department of Medical Sciences, University of Turin, Turin, Italy (Prof. L. Richiardi PhD)
- Netherlands Comprehensive Cancer Organisation (IKNL), Utrecht, Netherlands (O. Visser PhD MD)

**Contents:**

**Supplementary Appendix A.** Methods

**Supplementary Figure 1:** Age-standardised incidence rates of first-generation migrants in relation to birth-country and host-country incidence, by birth-country incidence categories, cancer site and sex in five European population-based cancer registries, 2013–17.
**Supplementary Figure 2A:** Cumulative risk ratios for first-generation migrants relative to the host population, by Human Development Index category of the country of birth, cancer site, and sex, in six European population-based cancer registries, 2013–17.
**Supplementary Figure 2B:** Cumulative risk ratios for first-generation migrants relative to the host population, by UN region of the country of birth, cancer site, and sex, in six European population-based cancer registries, 2013–17.
**Supplementary Figure 3:** Cumulative risk ratios for first-generation migrants relative to the host population by period, cancer site, sex, and registry, in five European population-based cancer registries 2003–22.
**Supplementary Figure 4: (A)** Age and sex structure of the population aged 20 years and over in percentage by sex and and **(B)** proportion of women in the population per age, in percentage, in Ukraine, among Ukrainian-born residents of Poland, and in the host Polish population, 2022–23.

**Supplementary Table 1A:** Data availability in Cancer RADAR: observation periods and data sources for cancer cases and person-years at risk, by cancer registry.
**Supplementary Table 1B:** Birth-country incidence categories: range of GLOBOCAN 2022 age-standardised incidence rates of the countries of birth in each category, by cancer site and sex.
**Supplementary Table 1C:** Countries of birth represented in each birth-country incidence category, by registry, sex, and cancer site, 2013–17.
**Supplementary Table 2:** Incident cancer cases and person-years in host and first-generation migrant populations, by cancer site and registry, 2013–17.
**Supplementary Table 3:** Change over time in the cumulative risk ratio for first-generation migrants relative to the host population, by cancer site, sex, and registry.
**Supplementary Table 4:** Data quality indicators by cancer registry, migration status, sex, and cancer site.

### Supplementary Appendix A. Methods

#### A1. Data sources and data-quality indicators

Cancer RADAR applied the data definitions and quality checks established by IARC's Cancer Incidence in Five Continents (CI5) series. Three data-quality indicators were collected for each contributing registry:

1. Percentage of cases registered from a death certificate only (DCO%), stratified by country of birth.
2. Percentage of microscopically verified cases (MV%), stratified by country of birth.
3. Percentage of cases for whom country of birth was missing.

The type of data and data availability for each registry are summarised in Supplementary Table 1A. Indicator values by registry, sex, cancer site, and migration status are given in Supplementary Table 4.

Registries identified first-generation migrants directly from registry records, with the exception of Belgium, where migrant status was obtained through linkage with census data held by the national statistical office (section A2).

#### A2. Belgium: census linkage and age truncation

In Belgium, first-generation migrant status was identified through linkage with the 2001 national census, restricted to individuals aged 18 years and over at the census baseline of 1 October 2001. Because this closed cohort was followed forward over time, younger age groups were progressively no longer observed in successive study periods.

Belgian cumulative risks were therefore calculated over a truncated age range, and the truncation window shifted upward across periods as the cohort aged:

| Study period | Age range for Belgian CRs |
|---|---|
| 2004–07 | 20–74 years |
| 2008–12 | 25–74 years |
| 2013–17 | 30–74 years |
| 2018–22 | 35–74 years |

In addition, because national Belgian Cancer Registry data are complete only from 2004 onwards, the first study period covers 2004–07 rather than 2003–07. Linkage to country of birth was performed only for the six site-specific cancers included in this study. Belgium therefore does not contribute to the all-cancers-combined analyses.

Belgian cumulative risks remain comparable with those of the other registries when interpreted over their specified age ranges, but underestimate full-range risk. Because the truncation window shifts upward across periods, Belgian trend estimates compare slightly different age ranges. Age-standardised incidence rates are not reported for Belgium, which is therefore excluded from comparisons on the ASR scale.

**Supplementary Figure 1:** Age-standardised incidence rates of first-generation migrants in relation to birth-country and host-country incidence, by birth-country incidence categories, cancer site and sex in five European population-based cancer registries, 2013–17.

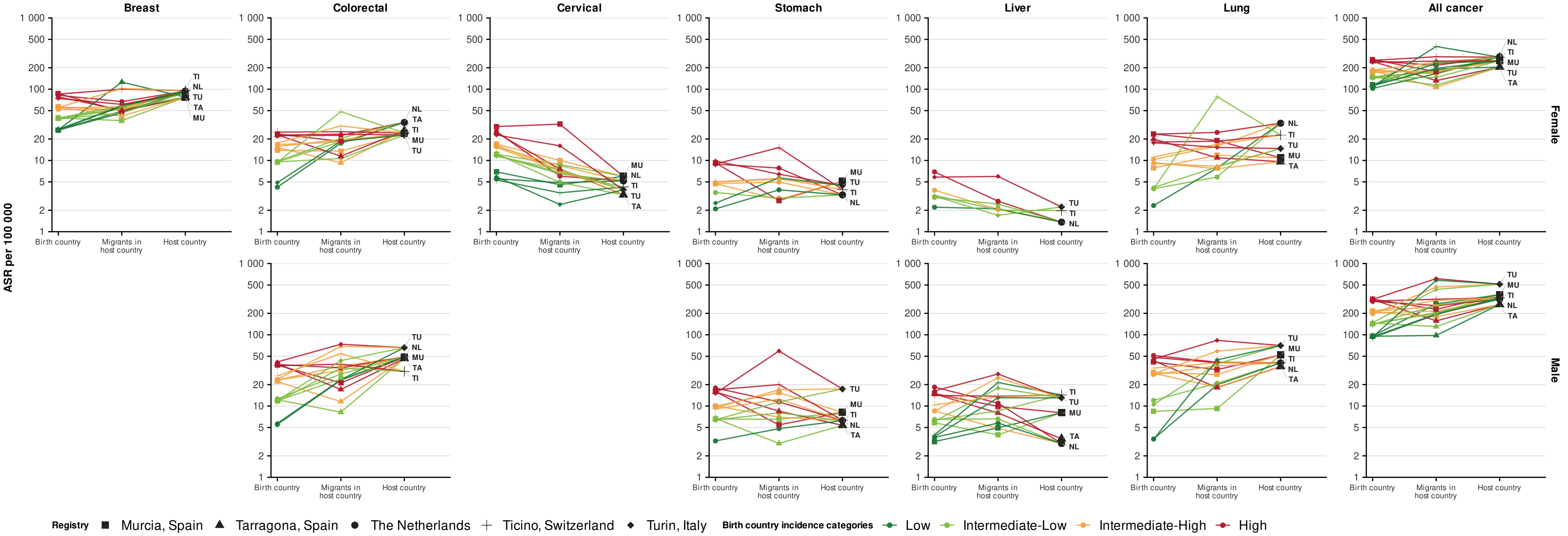


ASR = age-standardised incidence rate per 100 000, log scale. Each panel shows one cancer site; within each registry, four lines correspond to the four birth-country incidence categories, defined separately for each cancer site and sex from GLOBOCAN 2022 age-standardised incidence rates (Supplementary Table 1B), and each symbol marks one registry. The three categories on the X axis are **(1) birth-country** = birth country CR, a person-year-weighted average of the GLOBOCAN 2022 cumulative risks of the countries of birth in that category, over the same age range; **(2) migrants in host country** = the observed CR among first-generation migrants in the registry; and **(3) host country** = the observed CR in the host population. All panels share one Y axis range. All panels share one Y axis range. Strata with fewer than 5 cases are not shown. Belgium is not shown, because its age-truncated rates are not compared on the ASR scale.

**Supplementary Figure 2A:** Cumulative risk ratios for first-generation migrants relative to the host population, by Human Development Index category of the country of birth, cancer site, and sex, in six European population-based cancer registries, 2013–17.

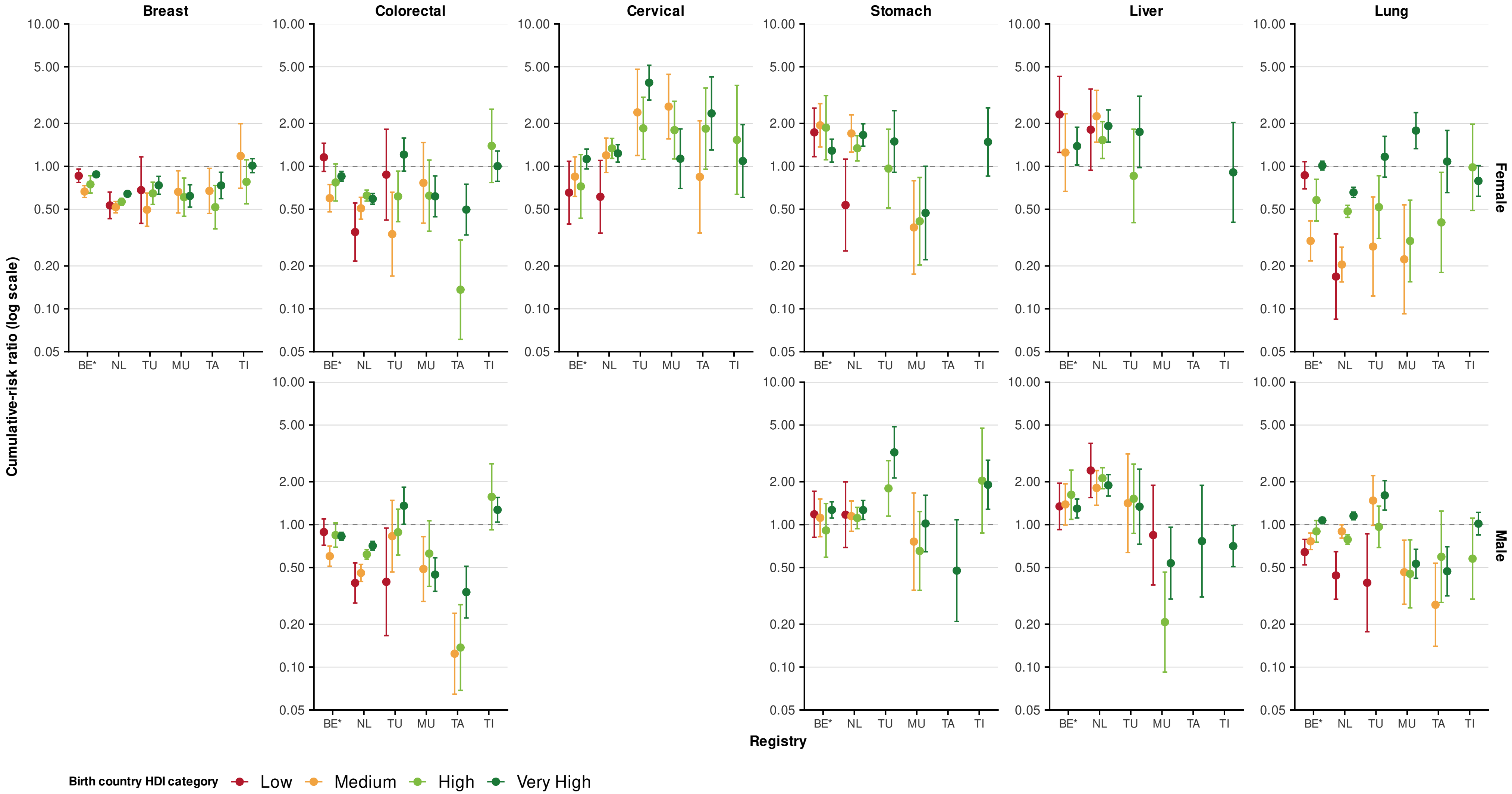


CRR = cumulative risk ratio (migrants vs host), based on cumulative risk from age 20 to 74 years; HDI = Human Development Index. BE = Belgium; NL = the Netherlands; TU = Turin, Italy; MU = Murcia, Spain; TA = Tarragona, Spain; TI = Ticino, Switzerland. Within each panel, registries are on the X axis and the four UNDP HDI 2023 categories of the country of birth (low, medium, high, very high) are shown as coloured points with 95% CI bars; the Y axis is on a log scale, and the dashed line marks a CRR of 1. Strata with fewer than 5 cases are not shown. Breast and cervical cancer are shown for females only. * In Belgium (BE*), first-generation migrant status was obtained through linkage with the 2001 national census, restricted to individuals aged ≥18 years at the census baseline (1 October 2001). Because this closed cohort was followed forward, the observable age range is incomplete and shifts towards older ages over time. Belgian CRs for 2013–17 are therefore age-truncated to ages 30–74 years. They remain comparable with those of the other registries but underestimate full-range risk.

**Supplementary Figure 2B:** Cumulative risk ratios for first-generation migrants relative to the host population, by UN region of the country of birth, cancer site, and sex, in six European population-based cancer registries, 2013–17.

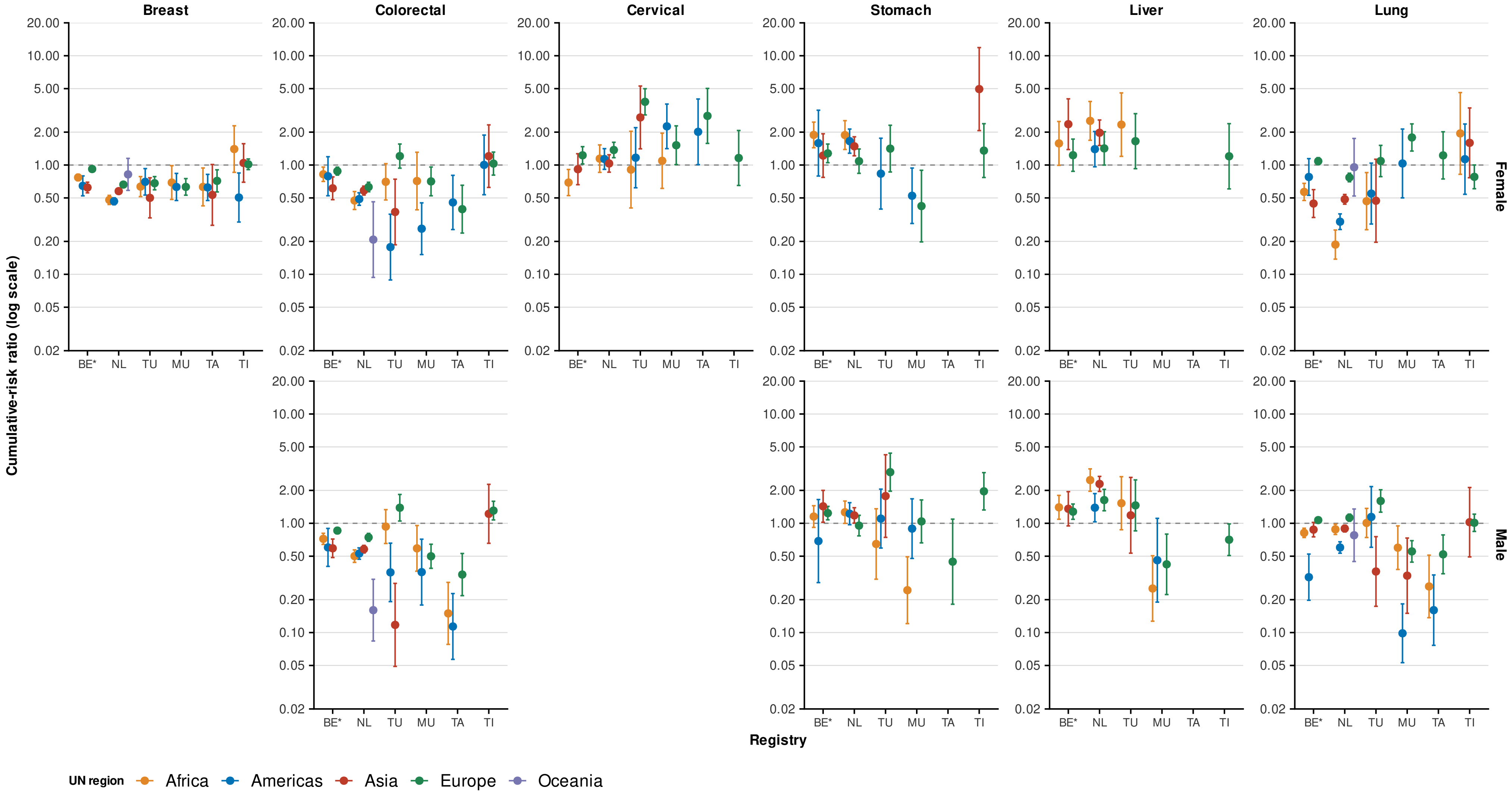


CRR = cumulative risk ratio (migrants vs host), based on cumulative risk from age 20 to 74 years; UN = United Nations. . BE = Belgium; NL = the Netherlands; TU = Turin, Italy; MU = Murcia, Spain; TA = Tarragona, Spain; TI = Ticino, Switzerland. Within each panel, registries are on the X axis and the five UN regions of the country of birth (Africa, Americas, Asia, Europe, Oceania) are shown as coloured points with 95% CI bars; the Y axis is on a log scale, and the dashed line marks a CRR of 1. Strata with fewer than 5 cases are not shown. Breast and cervical cancer are shown for females only. * In Belgium (BE*), first-generation migrant status was obtained through linkage with the 2001 national census, restricted to individuals aged ≥18 years at the census baseline (1 October 2001). Because this closed cohort was followed forward, the observable age range is incomplete and shifts towards older ages over time. Belgian CRs for 2013–17 are therefore age-truncated to ages 30–74 years. They remain comparable with those of the other registries but underestimate full-range risk.

**Supplementary Figure 3:** Cumulative risk ratios for first-generation migrants relative to the host population by period, cancer site, sex, and registry, in five European population-based cancer registries, 2003–22.

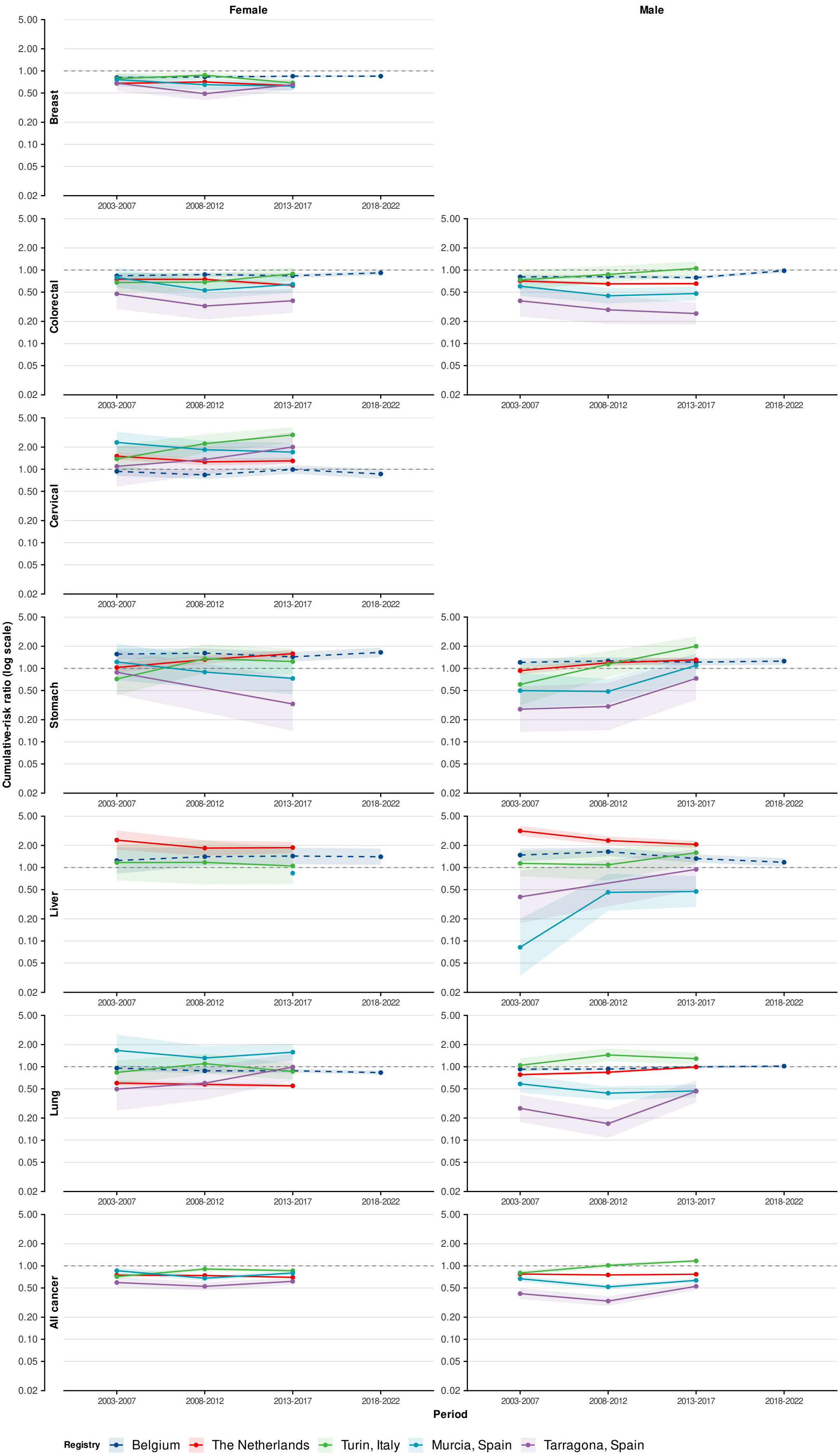

CRR = cumulative risk ratio, based on cumulative risk from age 20 to 74 years, log scale. Lines show the CRR in each period with 95% CI as shaded ribbons; the horizontal grey dashed line marks a CRR of 1, and all panels share one Y axis range. Only registries with at least two periods of data are shown: Ticino provided person-years for 2013–17 only, so Ticino is not included in this analysis. Percentage changes between periods, with 95% CIs, are in Supplementary Table 3.

Belgium (blue dashed line) is a closed census-based cohort: first-generation migrant status was obtained through linkage with the 1 October 2001 national census, restricted to individuals aged 18 years or older at that date, with national registry data complete from 2004. Its first period covers 2004–07, and its age range rises across periods (20–74 years in 2004–07, 25–74 in 2008–12, 30–74 in 2013–17, and 35–74 in 2018–22), so Belgian comparisons across periods span different age ranges.

**Interpretation:** CRRs comparing first-generation migrants with the host population are plotted for each study period (2003–07, 2008–12, 2013–17, and 2018–22 for Belgium), with one line per registry. A CRR above 1 indicates higher cumulative risk among migrants than in the host population, and the slope of each line indicates the direction and magnitude of change over time.

**Supplementary Figure 4: (A)** Age and sex structure of the population aged 20 years and over in percentage by sex and and **(B)** proportion of women in the population per age, in percentage, in Ukraine, among Ukrainian-born residents of Poland, and in the host Polish population, 2022–23.

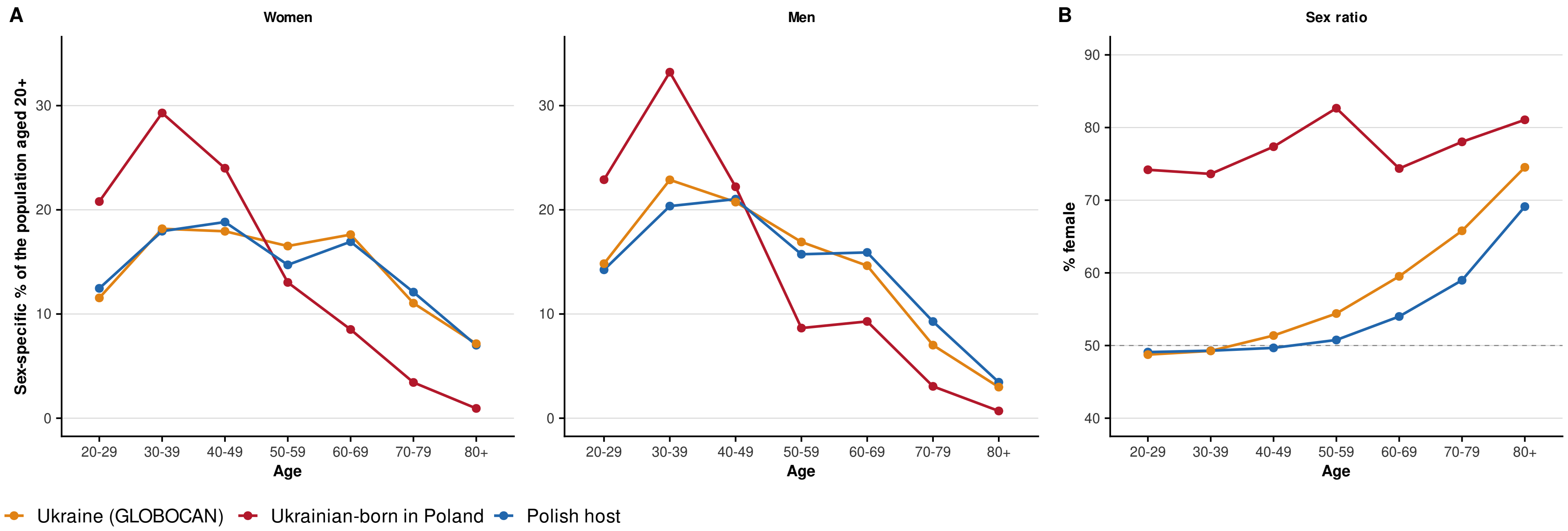


Lines show the percentage of each population in each 10-year age group, calculated separately for women and men. (A) Age distribution of women and of men. (B) Percentage of women in each age group; the dashed line marks equal numbers of women and men (50%). Among Ukrainian-born residents of Poland, 74% of women and 78% of men were younger than 50 years, compared with 49% and 56% in the host Polish population and 48% and 58% in Ukraine. Women made up 76% of the Ukrainian-born population (compared with 52% in the host Polish population and 55% in Ukraine).

**Supplementary Table 1A:** Data availability in Cancer RADAR: observation periods and data sources for cancer cases and person-years at risk, by cancer registry.

| Registry | Periods data available | Source of country-of-birth data for cancer cases | Source of person-year data | Other comments |
|---|---|---|---|---|
| Belgium (national) | 2004–07; 2008–12; 2013–17; 2018–22 | National census | National census | In Belgium, first-generation migrant status was identified through linkage with data from the national census of 2001 including individuals aged ≥18 years at the census baseline (1 October 2001). |
| Netherlands (national) | 2003–07; 2008–12; 2013–17; | Hospital medical records | National statistical office | |
| Turin, Italy | 2003–07; 2008–12; 2013–17; | National population register | National statistical office | |
| Murcia, Spain | 2003–07; 2008–12; 2013–17; | Regional population register | National statistical office | |
| Tarragona, Spain | 2003–07; 2008–12; 2013–17; | Health insurance register | Catalan statistical officeThe Statistical Institute of Catalonia | |
| Ticino, Switzerland | 2003–07; 2008–12; 2013–17; | National population register | National statistical office | For Ticino, person-year data were available only for 2013–17. Ticino was therefore excluded from the time-trend analyses. |
| Poland (national) | 2022–23 | National population register | National statistical office | Only data on host and Ukrainian population was collected. |

For all cancer registries, country-of-birth data for cancer cases was available directly within the registry. Belgium was the exception, where data linkage with external sources was necessary.

**Supplementary Table 1B:** Birth-country incidence categories: range of GLOBOCAN 2022 age-standardised incidence rates of the countries of birth in each category, by cancer site and sex.

| Cancer site | Sex | Low | Intermediate-low | Intermediate-high | High |
|---|---|---|---|---|---|
| Breast | Female | ≤ 30·98 | > 30·98 to 46·21 | > 46·21 to 63·12 | > 63·12 |
| Colorectal | Female | ≤ 3·23 | > 3·23 to 7·45 | > 7·45 to 13·44 | > 13·44 |
| Colorectal | Male | ≤ 4·15 | > 4·15 to 8·74 | > 8·74 to 17·56 | > 17·56 |
| Cervical | Female | ≤ 8·01 | > 8·01 to 13·75 | > 13·75 to 21·49 | > 21·49 |
| Stomach | Female | ≤ 3·11 | > 3·11 to 4·12 | > 4·12 to 6·41 | > 6·41 |
| Stomach | Male | ≤ 4·52 | > 4·52 to 7·37 | > 7·37 to 13·02 | > 13·02 |
| Liver | Female | ≤ 2·58 | > 2·58 to 3·44 | > 3·44 to 5·02 | > 5·02 |
| Liver | Male | ≤ 5·24 | > 5·24 to 6·95 | > 6·95 to 10·94 | > 10·94 |
| Lung | Female | ≤ 3·18 | > 3·18 to 6·02 | > 6·02 to 13·50 | > 13·50 |
| Lung | Male | ≤ 6·43 | > 6·43 to 16·98 | > 16·98 to 35·83 | > 35·83 |
| All cancer* | Female | ≤ 126·18 | > 126·18 to 155·72 | > 155·72 to 211·17 | > 211·17 |
| All cancer* | Male | ≤ 106·68 | > 106·68 to 162·00 | > 162·00 to 253·85 | > 253·85 |

Values are birth-country age-standardised incidence rates (ASRs) per 100 000 person-years, standardised to the World Standard Population, obtained from GLOBOCAN 2022. Quartile boundaries were defined separately for each cancer site and sex because countries occupy different relative positions across cancer types. Migrants were assigned to a birth-country risk group according to the ASR of their birth country for the cancer site under consideration.

* All cancer sites combined (C00–C97, excluding C44)

### Supplementary Table 1C: Countries of birth represented in each birth-country incidence category, by registry, sex, and cancer site, 2013–17.

| Registry | Sex | Cancer type | Burden category | Countries (N) | Total cases | Origin countries (cases) |
|---|---|---|---|---|---|---|
| Belgium | Female | Breast | Low | 8 | 459 | Democratic Republic of the Congo (357); Congo (25); Rwanda (24); Other, 5 countries (53) |
| Belgium | Female | Breast | Intermediate-Low | 10 | 118 | Tunisia (27); Other, 9 countries (91) |
| Belgium | Female | Breast | Intermediate-High | 12 | 845 | Morocco (413); Türkiye (236); Algeria (73); Philippines (38); Other, 8 countries (85) |
| Belgium | Female | Breast | High | 22 | 3284 | France (849); Italy (780); Netherlands (Kingdom of the) (455); Germany (426); Spain (178); Other, 17 countries (596) |
| Belgium | Female | Colorectal | Low | 3 | 130 | Democratic Republic of the Congo (113); Other, 2 countries (17) |
| Belgium | Female | Colorectal | Intermediate-Low | 4 | 131 | Morocco (104); Other, 3 countries (27) |
| Belgium | Female | Colorectal | Intermediate-High | 8 | 238 | Germany (134); Türkiye (53); Other, 6 countries (51) |
| Belgium | Female | Colorectal | High | 13 | 1122 | Italy (337); France (313); Netherlands (Kingdom of the) (203); Spain (84); Poland (49); Other, 8 countries (136) |
| Belgium | Female | Cervical | Low | 7 | 171 | France (53); Germany (29); Netherlands (Kingdom of the) (29); Italy (28); Other, 3 countries (32) |
| Belgium | Female | Cervical | Intermediate-Low | 3 | 50 | Morocco (30); Other, 2 countries (20) |
| Belgium | Female | Cervical | Intermediate-High | 1 | 5 | Other, 1 country (5) |
| Belgium | Female | Cervical | High | 2 | 26 | Other, 2 countries (26) |
| Belgium | Female | Stomach | Low | 1 | 48 | France (48) |
| Belgium | Female | Stomach | Intermediate-Low | 2 | 57 | Netherlands (Kingdom of the) (33); Democratic Republic of the Congo (24) |
| Belgium | Female | Stomach | Intermediate-High | 7 | 189 | Italy (79); Morocco (49); Spain (27); Other, 4 countries (34) |
| Belgium | Female | Stomach | High | 2 | 30 | Türkiye (20); Other, 1 country (10) |
| Belgium | Female | Liver | Low | 2 | 16 | Other, 2 countries (16) |
| Belgium | Female | Liver | Intermediate-Low | 3 | 56 | Italy (27); Other, 2 countries (29) |
| Belgium | Female | Liver | Intermediate-High | 1 | 10 | Other, 1 country (10) |
| Belgium | Female | Lung | Low | 3 | 106 | Democratic Republic of the Congo (94); Other, 2 countries (12) |
| Belgium | Female | Lung | Intermediate-Low | 2 | 43 | Morocco (38); Other, 1 country (5) |
| Belgium | Female | Lung | Intermediate-High | 6 | 74 | Türkiye (36); Other, 5 countries (38) |
| Belgium | Female | Lung | High | 14 | 1063 | France (281); Italy (214); Netherlands (Kingdom of the) (203); Germany (158); Spain (53); Other, 9 countries (154) |
| Belgium | Male | Colorectal | Low | 4 | 154 | Democratic Republic of the Congo (127); Other, 3 countries (27) |
| Belgium | Male | Colorectal | Intermediate-Low | 5 | 228 | Morocco (187); Other, 4 countries (41) |
| Belgium | Male | Colorectal | Intermediate-High | 6 | 314 | Germany (146); Türkiye (83); Algeria (57); Other, 3 countries (28) |
| Belgium | Male | Colorectal | High | 11 | 1301 | Italy (505); France (291); Netherlands (Kingdom of the) (237); Spain (93); Portugal (37); Other, 6 countries (138) |
| Belgium | Male | Stomach | Low | 1 | 22 | Democratic Republic of the Congo (22) |
| Belgium | Male | Stomach | Intermediate-Low | 5 | 170 | Morocco (63); France (54); Netherlands (Kingdom of the) (43); Other, 2 countries (10) |

| | | | | | | |
|---|---|---|---|---|---|---|
| Belgium | Male | Stomach | Intermediate-High | 7 | 232 | Italy (140); Spain (29); Germany (24); Other, 4 countries (39) |
| Belgium | Male | Stomach | High | 2 | 57 | Türkiye (44); Other, 1 country (13) |
| Belgium | Male | Liver | Low | 3 | 68 | Morocco (43); Other, 2 countries (25) |
| Belgium | Male | Liver | Intermediate-Low | 2 | 37 | Türkiye (24); Other, 1 country (13) |
| Belgium | Male | Liver | Intermediate-High | 3 | 137 | Italy (98); Democratic Republic of the Congo (24); Other, 1 country (15) |
| Belgium | Male | Liver | High | 5 | 93 | France (66); Other, 4 countries (27) |
| Belgium | Male | Lung | Low | 3 | 160 | Democratic Republic of the Congo (142); Other, 2 countries (18) |
| Belgium | Male | Lung | Intermediate-Low | 2 | 14 | Other, 2 countries (14) |
| Belgium | Male | Lung | Intermediate-High | 10 | 1147 | Italy (820); Germany (159); Algeria (72); United Kingdom of Great Britain and Northern Ireland (43); Other, 6 countries (53) |
| Belgium | Male | Lung | High | 12 | 1622 | France (457); Morocco (324); Netherlands (Kingdom of the) (277); Türkiye (206); Spain (122); Other, 7 countries (236) |
| The Netherlands | Female | Breast | Low | 5 | 166 | Iran (Islamic Republic of) (51); Afghanistan (46); India (36); Cabo Verde (25); Other, 1 country (8) |
| The Netherlands | Female | Breast | Intermediate-Low | 16 | 1065 | Indonesia (717); China (105); Syrian Arab Republic (42); Viet Nam (34); Thailand (30); Other, 11 countries (137) |
| The Netherlands | Female | Breast | Intermediate-High | 23 | 2006 | Suriname (686); Türkiye (522); Morocco (399); Iraq (80); Bulgaria (41); Other, 18 countries (278) |
| The Netherlands | Female | Breast | High | 29 | 1691 | Germany (469); Belgium (191); Poland (186); United Kingdom of Great Britain and Northern Ireland (174); France (74); Other, 24 countries (597) |
| The Netherlands | Female | Colorectal | Low | 4 | 42 | Other, 4 countries (42) |
| The Netherlands | Female | Colorectal | Intermediate-Low | 8 | 664 | Indonesia (499); Morocco (102); Other, 6 countries (63) |
| The Netherlands | Female | Colorectal | Intermediate-High | 9 | 479 | Germany (239); Türkiye (136); China (52); Other, 6 countries (52) |
| The Netherlands | Female | Colorectal | High | 22 | 669 | Suriname (273); Belgium (83); United Kingdom of Great Britain and Northern Ireland (68); Poland (59); Spain (30); Other, 17 countries (156) |
| The Netherlands | Female | Cervical | Low | 7 | 113 | Türkiye (48); Germany (29); Other, 5 countries (36) |
| The Netherlands | Female | Cervical | Intermediate-Low | 5 | 111 | Poland (49); Morocco (46); Other, 3 countries (16) |
| The Netherlands | Female | Cervical | Intermediate-High | 4 | 51 | Other, 4 countries (51) |
| The Netherlands | Female | Cervical | High | 3 | 127 | Suriname (76); Indonesia (40); Other, 1 country (11) |
| The Netherlands | Female | Stomach | Low | 4 | 126 | Suriname (81); Indonesia (30); Other, 2 countries (15) |
| The Netherlands | Female | Stomach | Intermediate-Low | 1 | 9 | Other, 1 country (9) |
| The Netherlands | Female | Stomach | Intermediate-High | 6 | 124 | Morocco (56); Germany (41); Other, 4 countries (27) |
| The Netherlands | Female | Stomach | High | 3 | 107 | Türkiye (83); Other, 2 countries (24) |
| The Netherlands | Female | Liver | Low | 3 | 41 | Germany (22); Other, 2 countries (19) |
| The Netherlands | Female | Liver | Intermediate-Low | 2 | 36 | Türkiye (29); Other, 1 country (7) |
| The Netherlands | Female | Liver | Intermediate-High | 1 | 34 | Indonesia (34) |
| The Netherlands | Female | Liver | High | 1 | 25 | Suriname (25) |
| The Netherlands | Female | Lung | Low | 1 | 6 | Other, 1 country (6) |

| | | | | | | |
|---|---|---|---|---|---|---|
| The Netherlands | Female | Lung | Intermediate-Low | 3 | 51 | Morocco (34); Other, 2 countries (17) |
| The Netherlands | Female | Lung | Intermediate-High | 14 | 765 | Indonesia (426); Suriname (166); Türkiye (88); Other, 11 countries (85) |
| The Netherlands | Female | Lung | High | 19 | 681 | Germany (266); Belgium (95); United Kingdom of Great Britain and Northern Ireland (72); Poland (63); China (31); Other, 14 countries (154) |
| The Netherlands | Female | All cancer | Low | 11 | 488 | Afghanistan (134); India (100); Cabo Verde (89); Pakistan (47); Sri Lanka (29); Other, 6 countries (89) |
| The Netherlands | Female | All cancer | Intermediate-Low | 26 | 7198 | Indonesia (2912); Suriname (2150); Morocco (1123); Iraq (209); Iran (Islamic Republic of) (133); Other, 21 countries (671) |
| The Netherlands | Female | All cancer | Intermediate-High | 25 | 2693 | Türkiye (1521); China (329); Bulgaria (120); South Africa (113); Philippines (111); Other, 20 countries (499) |
| The Netherlands | Female | All cancer | High | 35 | 5874 | Germany (1920); Belgium (678); Poland (652); United Kingdom of Great Britain and Northern Ireland (565); France (220); Other, 30 countries (1839) |
| The Netherlands | Male | Colorectal | Low | 9 | 102 | Cabo Verde (22); Other, 8 countries (80) |
| The Netherlands | Male | Colorectal | Intermediate-Low | 8 | 856 | Indonesia (558); Morocco (197); Iraq (28); Viet Nam (25); Other, 4 countries (48) |
| The Netherlands | Male | Colorectal | Intermediate-High | 8 | 921 | Suriname (324); Germany (257); Türkiye (242); China (49); Iran (Islamic Republic of) (22); Other, 3 countries (27) |
| The Netherlands | Male | Colorectal | High | 16 | 516 | United Kingdom of Great Britain and Northern Ireland (99); Italy (86); Belgium (79); Poland (51); Spain (49); Other, 11 countries (152) |
| The Netherlands | Male | Stomach | Low | 2 | 69 | Indonesia (59); Other, 1 country (10) |
| The Netherlands | Male | Stomach | Intermediate-Low | 6 | 228 | Suriname (111); Morocco (80); Other, 4 countries (37) |
| The Netherlands | Male | Stomach | Intermediate-High | 5 | 91 | Germany (44); Other, 4 countries (47) |
| The Netherlands | Male | Stomach | High | 6 | 175 | Türkiye (121); China (20); Other, 4 countries (34) |
| The Netherlands | Male | Liver | Low | 5 | 75 | Morocco (47); Other, 4 countries (28) |
| The Netherlands | Male | Liver | Intermediate-Low | 4 | 133 | Türkiye (70); Germany (28); United Kingdom of Great Britain and Northern Ireland (22); Other, 1 country (13) |
| The Netherlands | Male | Liver | Intermediate-High | 3 | 79 | Suriname (56); Other, 2 countries (23) |
| The Netherlands | Male | Liver | High | 7 | 152 | Indonesia (59); China (31); Egypt (22); Viet Nam (20); Other, 3 countries (20) |
| The Netherlands | Male | Lung | Low | 5 | 40 | Other, 5 countries (40) |
| The Netherlands | Male | Lung | Intermediate-Low | 6 | 75 | Iran (Islamic Republic of) (21); Other, 5 countries (54) |
| The Netherlands | Male | Lung | Intermediate-High | 19 | 1582 | Indonesia (545); Suriname (324); Germany (311); Italy (119); United Kingdom of Great Britain and Northern Ireland (113); Other, 14 countries (170) |
| The Netherlands | Male | Lung | High | 14 | 1223 | Türkiye (429); Morocco (375); Belgium (85); Spain (75); China (53); Other, 9 countries (206) |
| The Netherlands | Male | All cancer | Low | 19 | 890 | Cabo Verde (150); Afghanistan (138); Ghana (123); India (107); Somalia (101); Other, 14 countries (271) |
| The Netherlands | Male | All cancer | Intermediate-Low | 18 | 5854 | Indonesia (3186); Morocco (1860); Iraq (250); Iran (Islamic Republic of) (193); Syrian Arab Republic (139); Other, 13 countries (226) |
| The Netherlands | Male | All cancer | Intermediate-High | 27 | 3302 | Suriname (2228); China (312); Egypt (148); Viet Nam (106); Bulgaria (73); Other, 22 countries (435) |
| The Netherlands | Male | All cancer | High | 29 | 7736 | Türkiye (2118); Germany (1662); United Kingdom of Great Britain and Northern Ireland (749); Italy (603); Belgium (517); Other, 24 countries (2087) |
| Turin, Italy | Female | Breast | Low | 2 | 19 | Other, 2 countries (19) |
| Turin, Italy | Female | Breast | Intermediate-Low | 5 | 62 | Tunisia (26); Peru (21); Other, 3 countries (15) |

| | | | | | | |
|---|---|---|---|---|---|---|
| Turin, Italy | Female | Breast | Intermediate-High | 10 | 160 | Morocco (41); Albania (27); Republic of Moldova (24); Other, 7 countries (68) |
| Turin, Italy | Female | Breast | High | 9 | 307 | Romania (177); France (43); Germany (26); Other, 6 countries (61) |
| Turin, Italy | Female | Colorectal | Low | 1 | 5 | Other, 1 country (5) |
| Turin, Italy | Female | Colorectal | Intermediate-Low | 3 | 21 | Other, 3 countries (21) |
| Turin, Italy | Female | Colorectal | Intermediate-High | 4 | 32 | Other, 4 countries (32) |
| Turin, Italy | Female | Colorectal | High | 4 | 107 | Romania (63); France (32); Other, 2 countries (12) |
| Turin, Italy | Female | Cervical | Intermediate-Low | 2 | 10 | Other, 2 countries (10) |
| Turin, Italy | Female | Cervical | High | 1 | 69 | Romania (69) |
| Turin, Italy | Female | Stomach | Low | 1 | 6 | Other, 1 country (6) |
| Turin, Italy | Female | Stomach | Intermediate-High | 1 | 19 | Other, 1 country (19) |
| Turin, Italy | Female | Liver | High | 1 | 9 | Other, 1 country (9) |
| Turin, Italy | Female | Lung | Intermediate-Low | 1 | 7 | Other, 1 country (7) |
| Turin, Italy | Female | Lung | High | 3 | 48 | Romania (24); Other, 2 countries (24) |
| Turin, Italy | Female | All cancer | Low | 2 | 147 | Tunisia (100); Libya (47) |
| Turin, Italy | Female | All cancer | Intermediate-Low | 7 | 235 | Morocco (120); Albania (66); Other, 5 countries (49) |
| Turin, Italy | Female | All cancer | Intermediate-High | 15 | 378 | Peru (74); Republic of Moldova (64); Argentina (41); Philippines (37); Brazil (34); Other, 10 countries (128) |
| Turin, Italy | Female | All cancer | High | 18 | 1149 | Romania (637); France (191); Germany (70); Serbia (47); Switzerland (34); Other, 13 countries (170) |
| Turin, Italy | Male | Colorectal | Intermediate-Low | 2 | 36 | Tunisia (21); Other, 1 country (15) |
| Turin, Italy | Male | Colorectal | Intermediate-High | 1 | 15 | Other, 1 country (15) |
| Turin, Italy | Male | Colorectal | High | 3 | 68 | Romania (35); France (23); Other, 1 country (10) |
| Turin, Italy | Male | Stomach | Intermediate-Low | 1 | 7 | Other, 1 country (7) |
| Turin, Italy | Male | Stomach | High | 2 | 24 | Other, 2 countries (24) |
| Turin, Italy | Male | Liver | High | 3 | 20 | Other, 3 countries (20) |
| Turin, Italy | Male | Lung | Intermediate-High | 2 | 17 | Other, 2 countries (17) |
| Turin, Italy | Male | Lung | High | 6 | 132 | Romania (46); Tunisia (26); Morocco (25); Other, 3 countries (35) |
| Turin, Italy | Male | All cancer | Low | 3 | 42 | Ethiopia (27); Other, 2 countries (15) |
| Turin, Italy | Male | All cancer | Intermediate-Low | 8 | 409 | Tunisia (141); Morocco (123); Libya (89); Peru (26); Other, 4 countries (30) |
| Turin, Italy | Male | All cancer | Intermediate-High | 8 | 201 | Albania (64); Argentina (37); Egypt (33); China (27); Other, 4 countries (40) |
| Turin, Italy | Male | All cancer | High | 12 | 659 | Romania (317); France (142); Serbia (59); Belgium (27); Germany (23); Other, 7 countries (91) |
| Murcia, Spain | Female | Breast | Low | 1 | 12 | Other, 1 country (12) |
| Murcia, Spain | Female | Breast | Intermediate-Low | 2 | 37 | Ecuador (25); Other, 1 country (12) |
| Murcia, Spain | Female | Breast | Intermediate-High | 4 | 81 | Morocco (51); Other, 3 countries (30) |
| Murcia, Spain | Female | Breast | High | 7 | 128 | United Kingdom of Great Britain and Northern Ireland (54); France (24); Other, 5 countries (50) |

| | | | | | | |
|---|---|---|---|---|---|---|
| Murcia, Spain | Female | Colorectal | Intermediate-Low | 1 | 16 | Other, 1 country (16) |
| Murcia, Spain | Female | Colorectal | Intermediate-High | 2 | 14 | Other, 2 countries (14) |
| Murcia, Spain | Female | Colorectal | High | 3 | 37 | United Kingdom of Great Britain and Northern Ireland (22); Other, 2 countries (15) |
| Murcia, Spain | Female | Cervical | Intermediate-Low | 2 | 19 | Other, 2 countries (19) |
| Murcia, Spain | Female | Cervical | Intermediate-High | 2 | 16 | Other, 2 countries (16) |
| Murcia, Spain | Female | Cervical | High | 2 | 21 | Other, 2 countries (21) |
| Murcia, Spain | Female | Lung | High | 1 | 44 | United Kingdom of Great Britain and Northern Ireland (44) |
| Murcia, Spain | Female | All cancer | Low | 1 | 6 | Other, 1 country (6) |
| Murcia, Spain | Female | All cancer | Intermediate-Low | 4 | 243 | Morocco (172); Bolivia (Plurinational State of) (54); Other, 2 countries (17) |
| Murcia, Spain | Female | All cancer | Intermediate-High | 11 | 347 | Ecuador (123); Ukraine (53); Colombia (45); Bulgaria (38); Argentina (27); Other, 6 countries (61) |
| Murcia, Spain | Female | All cancer | High | 12 | 493 | United Kingdom of Great Britain and Northern Ireland (275); France (72); Romania (41); Germany (39); Other, 8 countries (66) |
| Murcia, Spain | Male | Colorectal | Intermediate-Low | 2 | 34 | Morocco (29); Other, 1 country (5) |
| Murcia, Spain | Male | Colorectal | Intermediate-High | 2 | 16 | Other, 2 countries (16) |
| Murcia, Spain | Male | Colorectal | High | 2 | 49 | United Kingdom of Great Britain and Northern Ireland (39); Other, 1 country (10) |
| Murcia, Spain | Male | Stomach | Intermediate-Low | 2 | 23 | Other, 2 countries (23) |
| Murcia, Spain | Male | Stomach | High | 1 | 5 | Other, 1 country (5) |
| Murcia, Spain | Male | Liver | Intermediate-Low | 1 | 8 | Other, 1 country (8) |
| Murcia, Spain | Male | Lung | Intermediate-High | 2 | 62 | United Kingdom of Great Britain and Northern Ireland (55); Other, 1 country (7) |
| Murcia, Spain | Male | Lung | High | 5 | 55 | Morocco (27); Other, 4 countries (28) |
| Murcia, Spain | Male | All cancer | Low | 2 | 15 | Other, 2 countries (15) |
| Murcia, Spain | Male | All cancer | Intermediate-Low | 5 | 372 | Morocco (239); Ecuador (91); Algeria (21); Other, 2 countries (21) |
| Murcia, Spain | Male | All cancer | Intermediate-High | 9 | 161 | Ukraine (38); Bulgaria (30); Argentina (27); Other, 6 countries (66) |
| Murcia, Spain | Male | All cancer | High | 12 | 705 | United Kingdom of Great Britain and Northern Ireland (444); France (84); Germany (41); Romania (36); Portugal (26); Other, 7 countries (74) |
| Tarragona, Spain | Female | Breast | Low | 1 | 5 | Other, 1 country (5) |
| Tarragona, Spain | Female | Breast | Intermediate-Low | 3 | 17 | Other, 3 countries (17) |
| Tarragona, Spain | Female | Breast | Intermediate-High | 4 | 58 | Morocco (34); Other, 3 countries (24) |
| Tarragona, Spain | Female | Breast | High | 6 | 74 | Romania (21); Other, 5 countries (53) |
| Tarragona, Spain | Female | Colorectal | High | 2 | 10 | Other, 2 countries (10) |
| Tarragona, Spain | Female | Cervical | High | 1 | 7 | Other, 1 country (7) |
| Tarragona, Spain | Female | All cancer | Intermediate-Low | 3 | 86 | Morocco (71); Other, 2 countries (15) |
| Tarragona, Spain | Female | All cancer | Intermediate-High | 12 | 146 | Argentina (29); Colombia (28); Other, 10 countries (89) |
| Tarragona, Spain | Female | All cancer | High | 11 | 211 | Romania (64); France (38); United Kingdom of Great Britain and Northern Ireland (29); Germany (21); Other, 7 countries (59) |

| | | | | | | |
|---|---|---|---|---|---|---|
| Tarragona, Spain | Male | Colorectal | Intermediate-Low | 1 | 6 | Other, 1 country (6) |
| Tarragona, Spain | Male | Colorectal | High | 2 | 15 | Other, 2 countries (15) |
| Tarragona, Spain | Male | Lung | Intermediate-High | 2 | 10 | Other, 2 countries (10) |
| Tarragona, Spain | Male | Lung | High | 2 | 21 | Other, 2 countries (21) |
| Tarragona, Spain | Male | All cancer | Low | 1 | 8 | Other, 1 country (8) |
| Tarragona, Spain | Male | All cancer | Intermediate-Low | 3 | 84 | Morocco (69); Other, 2 countries (15) |
| Tarragona, Spain | Male | All cancer | Intermediate-High | 7 | 80 | Argentina (24); Other, 6 countries (56) |
| Tarragona, Spain | Male | All cancer | High | 13 | 226 | Romania (54); United Kingdom of Great Britain and Northern Ireland (30); Germany (27); Italy (25); France (21); Other, 8 countries (69) |
| Ticino, Switzerland | Female | Breast | Intermediate-High | 6 | 57 | Other, 6 countries (57) |
| Ticino, Switzerland | Female | Breast | High | 11 | 542 | Italy (395); Germany (52); Other, 9 countries (95) |
| Ticino, Switzerland | Female | Colorectal | Intermediate-High | 3 | 37 | Germany (24); Other, 2 countries (13) |
| Ticino, Switzerland | Female | Colorectal | High | 1 | 151 | Italy (151) |
| Ticino, Switzerland | Female | Cervical | Low | 1 | 12 | Other, 1 country (12) |
| Ticino, Switzerland | Female | Stomach | Intermediate-High | 1 | 44 | Italy (44) |
| Ticino, Switzerland | Female | Liver | Intermediate-Low | 1 | 18 | Other, 1 country (18) |
| Ticino, Switzerland | Female | Lung | High | 4 | 122 | Italy (103); Other, 3 countries (19) |
| Ticino, Switzerland | Female | All cancer | Low | 2 | 12 | Other, 2 countries (12) |
| Ticino, Switzerland | Female | All cancer | Intermediate-Low | 3 | 30 | Other, 3 countries (30) |
| Ticino, Switzerland | Female | All cancer | Intermediate-High | 11 | 123 | Bosnia and Herzegovina (29); Türkiye (25); Brazil (20); Other, 8 countries (49) |
| Ticino, Switzerland | Female | All cancer | High | 22 | 1799 | Italy (1246); Germany (146); France (55); Serbia (53); Portugal (52); Other, 17 countries (247) |
| Ticino, Switzerland | Male | Colorectal | Intermediate-High | 3 | 38 | Other, 3 countries (38) |
| Ticino, Switzerland | Male | Colorectal | High | 4 | 249 | Italy (224); Other, 3 countries (25) |
| Ticino, Switzerland | Male | Stomach | Intermediate-High | 1 | 78 | Italy (78) |
| Ticino, Switzerland | Male | Stomach | High | 1 | 5 | Other, 1 country (5) |
| Ticino, Switzerland | Male | Liver | Intermediate-High | 1 | 73 | Italy (73) |
| Ticino, Switzerland | Male | Lung | Intermediate-High | 2 | 282 | Italy (272); Other, 1 country (10) |
| Ticino, Switzerland | Male | Lung | High | 3 | 18 | Other, 3 countries (18) |
| Ticino, Switzerland | Male | All cancer | Intermediate-High | 6 | 88 | Bosnia and Herzegovina (43); Other, 5 countries (45) |
| Ticino, Switzerland | Male | All cancer | High | 17 | 2185 | Italy (1756); Germany (94); Portugal (57); Serbia (52); France (48); Other, 12 countries (178) |

Note: Country counts below 5 are suppressed by the registries at source, so the cell totals here are lower than the migrant totals in Supplementary Table 2.

**Supplementary Table 2: Incident cancer cases and person-years in host and first-generation migrant populations, by cancer site and registry, 2013–17.**

| Registry | Cancer type / population | Women | | | Men | | |
|---|---|---|---|---|---|---|---|
| | | Host pop. | Migrant pop. | | Host pop. | Migrant pop. | |
| | | N | N | Prop. (%) Migrants | N | N | Prop. (%) Migrants |
| Belgium | Population (person-years) | 15 168 979 | 1 907 689 | 11·2 | 14 058 188 | 1 721 599 | 10·9 |
| | Breast | 46 411 | 4958 | 9·7 | n/a | n/a | n/a |
| | Colorectal | 16 393 | 1737 | 9·6 | 21 145 | 2136 | 9·2 |
| | Cervical | 2453 | 309 | 11·2 | n/a | n/a | n/a |
| | Stomach | 2198 | 375 | 14·6 | 3769 | 549 | 12·7 |
| | Liver | 611 | 106 | 14·8 | 2112 | 385 | 15·4 |
| | Lung | 11 494 | 1393 | 10·8 | 23 922 | 3095 | 11·5 |
| | All cancer | n/a | n/a | n/a | n/a | n/a | n/a |
| The Netherlands | Population (person-years) | 37 425 100 | 5 329 202 | 12·5 | 37 130 568 | 4 886 102 | 11·6 |
| | Breast | 63 567 | 5599 | 8·1 | n/a | n/a | n/a |
| | Colorectal | 30 266 | 2150 | 6·6 | 39 359 | 2743 | 6·5 |
| | Cervical | 2584 | 585 | 18·5 | n/a | n/a | n/a |
| | Stomach | 3040 | 479 | 13·6 | 5180 | 705 | 12·0 |
| | Liver | 1065 | 220 | 17·1 | 2338 | 559 | 19·3 |
| | Lung | 26 161 | 1751 | 6·3 | 33 632 | 3295 | 8·9 |
| | All cancer | 214 487 | 18 170 | 7·8 | 245 054 | 19 613 | 7·4 |
| Turin, Italy | Population (person-years) | 5 101 196 | 684 297 | 11·8 | 4 455 758 | 549 524 | 11·0 |
| | Breast | 9707 | 625 | 6·0 | n/a | n/a | n/a |
| | Colorectal | 3819 | 201 | 5·0 | 4832 | 169 | 3·4 |
| | Cervical | 358 | 110 | 23·5 | n/a | n/a | n/a |
| | Stomach | 778 | 48 | 5·8 | 1234 | 62 | 4·8 |
| | Liver | 387 | 27 | 6·5 | 996 | 57 | 5·4 |
| | Lung | 2156 | 113 | 5·0 | 5433 | 202 | 3·6 |
| | All cancer | 30 901 | 2027 | 6·2 | 37 972 | 1471 | 3·7 |
| Murcia, Spain | Population (person-years) | 3 135 145 | 525 172 | 14·3 | 3 068 187 | 612 771 | 16·6 |
| | Breast | 3833 | 309 | 7·5 | n/a | n/a | n/a |
| | Colorectal | 1642 | 103 | 5·9 | 2753 | 138 | 4·8 |
| | Cervical | 268 | 80 | 23·0 | n/a | n/a | n/a |

| | | | | | | | |
|---|---|---|---|---|---|---|---|
| | Stomach | 368 | 24 | 6·1 | 490 | 53 | 9·8 |
| | Liver | 73 | 6 | 7·6 | 411 | 35 | 7·8 |
| | Lung | 598 | 86 | 12·6 | 2775 | 158 | 5·4 |
| | All cancer | 13 750 | 1188 | 8·0 | 19 121 | 1350 | 6·6 |
| Tarragona, Spain | Population (person-years) | 1 656 379 | 335 715 | 16·9 | 1 639 377 | 357 326 | 17·9 |
| | Breast | 2221 | 209 | 8·6 | n/a | n/a | n/a |
| | Colorectal | 1017 | 35 | 3·3 | 1673 | 50 | 2·9 |
| | Cervical | 81 | 31 | 27·7 | n/a | n/a | n/a |
| | Stomach | 119 | 6 | 4·8 | 205 | 13 | 6·0 |
| | Liver | 27 | n/a | n/a | 112 | 13 | 10·4 |
| | Lung | 311 | 28 | 8·3 | 1251 | 56 | 4·3 |
| | All cancer | 6462 | 482 | 6·9 | 8892 | 441 | 4·7 |
| Ticino, Switzerland | Population (person-years) | 583 864 | 315 588 | 35·1 | 546 830 | 306 975 | 36·0 |
| | Breast | 1081 | 682 | 38·7 | n/a | n/a | n/a |
| | Colorectal | 371 | 249 | 40·2 | 373 | 332 | 47·1 |
| | Cervical | 43 | 28 | 39·4 | n/a | n/a | n/a |
| | Stomach | 65 | 63 | 49·2 | 82 | 106 | 56·4 |
| | Liver | 35 | 23 | 39·7 | 163 | 96 | 37·1 |
| | Lung | 304 | 172 | 36·1 | 475 | 346 | 42·1 |
| | All cancer | 3385 | 2098 | 38·3 | 3607 | 2420 | 40·2 |

Observed incident cancer cases (N) and person-years in the host (non-migrant) population and first-generation migrants, totals over five years, not annual averages. Prop. (%) = [pyo migrants / (pyo host + pyo migrants)] x 100. The Population row reports person-years; cancer rows report case counts. n/a = count suppressed or unavailable.

**Note:** Case counts are not directly comparable across tables because of differences in the level of aggregation. When sharing data, registries withhold any cell containing e.g. fewer than five cases before aggregated outputs are returned. Consequently, the finer the breakdown, the greater the number of cells withheld and the lower the resulting total. Totals for all migrants combined are reported directly by each registry rather than reconstructed by summing smaller groups and are therefore complete. In contrast, totals reconstructed from smaller groups may be lower because cases in withheld cells are excluded (**Supplementary Table 1C**).

**For example,** this table reports 309 cases of cervical cancer among females with a migration background in Belgium in 2013–17. Of these, 16 have an unknown country of birth, leaving 293 cases distributed across the birth-country incidence categories. While, supplementary Table 1C provides a more detailed breakdown, listing the individual countries within each category. Because country-specific cells containing fewer than five cases are withheld, the cases listed in **Supplementary Table 1C** sum to 252. The difference of 41 cases represents cases from countries for which fewer than five cases were recorded. Thus, cases in withheld cells are absent only from totals reconstructed by summing smaller groups. They remain included in the directly reported all-migrant totals and in categories that were defined a-priori (i.e. birth-country incidence categories).

**Supplementary Table 3:** Change over time in the cumulative risk ratio for first-generation migrants relative to the host population, by cancer site, sex, and registry.

| Registry | Sex | Cancer type | % change 2003–07 to 2013–17 (95% CI) | % change 2008–12 to 2013–17 (95% CI) |
|---|---|---|---|---|
| Belgium* | Female | Breast | 4·2 (−0·8 to 9·4) | 2·3 (−2·3 to 7·1) |
| The Netherlands | Female | Breast | −7·2 (−11·5 to −2·6) | −11·3 (−15·2 to −7·3) |
| Turin, Italy | Female | Breast | −12·6 (−26·9 to 4·5) | −21·5 (−33·2 to −7·8) |
| Murcia, Spain | Female | Breast | −18·3 (−36·8 to 5·5) | −4·0 (−22·5 to 18·8) |
| Tarragona, Spain | Female | Breast | −4·6 (−29·4 to 28·9) | 32·6 (1·7–73·0) |
| Belgium* | Female | Colorectal | 0·3 (−9·7 to 11·5) | −3·7 (−12·5 to 6·1) |
| The Netherlands | Female | Colorectal | −17·3 (−24·3 to −9·7) | −17·1 (−23·6 to −10·0) |
| Turin, Italy | Female | Colorectal | 31·1 (-9·8, 90·6) | 29·6 (-6·8, 80·3) |
| Murcia, Spain | Female | Colorectal | −19·2 (−46·2 to 21·3) | 20·8 (−17·6 to 77·1) |
| Tarragona, Spain | Female | Colorectal | −19·5 (−56·1 to 47·5) | 18·1 (−32·8 to 107·4) |
| Belgium* | Male | Colorectal | −2·2 (−10·5 to 6·8) | −3·0 (−10·4 to 5·1) |
| The Netherlands | Male | Colorectal | −8·1 (−14·9 to −0·7) | 0·8 (−6·1 to 8·2) |
| Turin, Italy | Male | Colorectal | 43·7 (−0·8 to 108·2) | 21·1 (−13·8 to 70·3) |
| Murcia, Spain | Male | Colorectal | −20·5 (−45·1 to 15·1) | 7·2 (−22·3 to 47·8) |
| Tarragona, Spain | Male | Colorectal | −33·0 (−63·3 to 22·4) | −11·2 (−49·3 to 55·7) |
| Belgium* | Female | Cervical | 6·1 (−12·5 to 28·7) | 18·4 (−2·1 to 43·4) |
| The Netherlands | Female | Cervical | −13·9 (−26·0 to 0·2) | 3·0 (−11·0 to 19·1) |
| Turin, Italy | Female | Cervical | 112·7 (29·7–248·9) | 31·5 (−10·4 to 93·2) |
| Murcia, Spain | Female | Cervical | −26·2 (−52·4 to 14·3) | −7·3 (−38·1 to 38·8) |
| Tarragona, Spain | Female | Cervical | 83·1 (−17·1 to 304·2) | 48·1 (−24·2 to 189·3) |
| Belgium* | Female | Stomach | −8·1 (−27·0 to 15·7) | −11·0 (−28·0 to 10·0) |
| The Netherlands | Female | Stomach | 54·3 (26·7–87·9) | 20·4 (0·7–44·0) |
| Turin, Italy | Female | Stomach | 72·7 (−9·7 to 230·2) | −8·7 (−50·3 to 67·7) |
| Murcia, Spain | Female | Stomach | −40·5 (−72·1 to 27·3) | −18·0 (−59·4 to 65·5) |
| Tarragona, Spain | Female | Stomach | −63·0 (−87·5 to 9·9) | n/a |
| Belgium* | Male | Stomach | 0·8 (−15·2 to 19·9) | −3·7 (−17·9 to 13·0) |
| The Netherlands | Male | Stomach | 40·2 (20·7–63·0) | 9·5 (−4·7 to 25·9) |
| Turin, Italy | Male | Stomach | 231·9 (61·2–583·2) | 75·8 (5·2–193·5) |
| Murcia, Spain | Male | Stomach | 120·5 (13·1–329·9) | 126·3 (31·6–289·4) |

| | | | | |
|---|---|---|---|---|
| Tarragona, Spain | Male | Stomach | 162·4 (−0·6 to 593·1) | 140·9 (−11·3 to 553·9) |
| Belgium* | Female | Liver | 15·0 (−29·5 to 87·7) | 2·0 (−30·6 to 50·0) |
| The Netherlands | Female | Liver | −21·0 (−44·3 to 12·2) | 1·6 (−24·8 to 37·3) |
| Turin, Italy | Female | Liver | −10·2 (−59·3 to 98·1) | −10·7 (−63·2 to 116·9) |
| Murcia, Spain | Female | Liver | n/a | n/a |
| Tarragona, Spain | Female | Liver | n/a | n/a |
| Belgium* | Male | Liver | −10·4 (−29·6 to 14·1) | −19·4 (−33·4 to −2·5) |
| The Netherlands | Male | Liver | −34·6 (−46·2 to −20·6) | −11·5 (−25·4 to 5·1) |
| Turin, Italy | Male | Liver | 39·8 (−18·6 to 140·1) | 46·3 (−20·2 to 168·1) |
| Murcia, Spain | Male | Liver | 475·0 (108·6–1484·6) | 2·7 (−51·7 to 118·5) |
| Tarragona, Spain | Male | Liver | 137·1 (−16·0 to 569·6) | n/a |
| Belgium* | Female | Lung | −8·0 (−17·9 to 3·0) | 0·2 (−9·2 to 10·6) |
| The Netherlands | Female | Lung | −8·3 (−16·9 to 1·1) | −4·4 (−12·6 to 4·4) |
| Turin, Italy | Female | Lung | 3·0 (−34·6 to 62·2) | −21·9 (−47·7 to 16·8) |
| Murcia, Spain | Female | Lung | −5·4 (−46·0 to 66·0) | 19·9 (−24·7 to 91·0) |
| Tarragona, Spain | Female | Lung | 99·7 (−9·6 to 341·3) | 65·6 (−15·3 to 223·9) |
| Belgium* | Male | Lung | 7·5 (0·3–15·2) | 7·1 (0·4–14·2) |
| The Netherlands | Male | Lung | 27·1 (18·9–35·8) | 17·8 (10·6–25·4) |
| Turin, Italy | Male | Lung | 23·2 (−7·6 to 64·1) | −11·0 (−32·2 to 16·8) |
| Murcia, Spain | Male | Lung | −19·7 (−43·1 to 13·3) | 7·5 (−19·8 to 43·9) |
| Tarragona, Spain | Male | Lung | 70·5 (−1·1 to 193·9) | 175·8 (57·2–384·0) |
| Belgium* | Female | All cancer | n/a | n/a |
| The Netherlands | Female | All cancer | −6·7 (−9·1 to −4·3) | −5·6 (−7·9 to −3·3) |
| Turin, Italy | Female | All cancer | 20·4 (8·6–33·5) | −5·1 (−13·2 to 3·8) |
| Murcia, Spain | Female | All cancer | −7·2 (−18·1 to 5·2) | 17·4 (5·6–30·6) |
| Tarragona, Spain | Female | All cancer | 3·8 (−15·0 to 26·7) | 17·0 (−1·4 to 38·8) |
| Belgium* | Male | All cancer | n/a | n/a |
| The Netherlands | Male | All cancer | −0·6 (−3·0 to 1·8) | 2·2 (−0·1 to 4·5) |
| Turin, Italy | Male | All cancer | 45·9 (32·2–61·1) | 15·2 (5·0–26·3) |
| Murcia, Spain | Male | All cancer | −5·1 (−15·3 to 6·4) | 22·4 (11·1–34·9) |
| Tarragona, Spain | Male | All cancer | 25·7 (2·1–54·7) | 58·9 (31·6–92·0) |

CRR = cumulative risk ratio, based on cumulative risk from age 20 to 74 years (first-generation migrants vs host), with 95% CI. Percentage change = ((CRR 2013–17 / CRR in the earlier period) − 1) × 100; positive values mean the ratio was higher in 2013–17. Its 95% CI is derived from the ratio of the two periods' CRRs on the log scale, treating the periods as independent. **The direction of a change must be read together with the CRR in each period, shown in Supplementary Figure 3.** Strata need at least 5 cases in both periods, otherwise we reported n/a. Belgian estimates cover 20–74 years in 2004–07, 25–74 in 2008–12, and 30–74 in 2013–17, so its comparisons span different age ranges.

**Supplementary Table 4:** Data quality indicators by cancer registry, migration status, sex, and cancer site.

**Interpretation:** Death certificate only (DCO) proportions were low (<1% in most strata) and microscopic verification (MV) high (generally >90%), and both were comparable between first-generation migrants and host populations across registries and cancer sites. MV was lower for liver cancer (17–83%), consistent with its often radiological diagnosis. Missingness of birth-country information was generally low or absent, except in Tarragona, where it reached up to a third of cases for some sites (stomach [34%, males], cervical [26%], liver, and lung) but was considerably lower for breast cancer (9%) and across all cancers combined (17% in women, 22% in men).

| Registry | Sex | Cancer type | Migrant population | | | | Host population | | | | | |
|---|---|---|---|---|---|---|---|---|---|---|---|---|
| | | | DCO (N) | % DCO | MV (N) | % MV | DCO (N) | % DCO | MV (N) | % MV | Missing birth country (N) | % missing birth country |
| The Netherlands | Female | Breast | 0 | 0·0 | 5590 | 99·8 | 0 | 0·0 | 63472 | 99·9 | 3788 | 5·2 |
| The Netherlands | Female | Colorectal | 0 | 0·0 | 2099 | 97·6 | 0 | 0·0 | 29432 | 97·2 | 563 | 1·7 |
| The Netherlands | Female | Cervical | 0 | 0·0 | 581 | 99·3 | 0 | 0·0 | 2576 | 99·7 | 549 | 14·8 |
| The Netherlands | Female | Stomach | 0 | 0·0 | 474 | 99·0 | 0 | 0·0 | 2970 | 97·7 | 43 | 1·2 |
| The Netherlands | Female | Liver | 0 | 0·0 | 138 | 62·7 | 0 | 0·0 | 749 | 70·3 | < 5 | < 0·4 |
| The Netherlands | Female | Lung | 0 | 0·0 | 1532 | 87·5 | 0 | 0·0 | 22662 | 86·6 | 119 | 0·4 |
| The Netherlands | Female | All cancer | 0 | 0·0 | 17386 | 95·7 | 0 | 0·0 | 204091 | 95·2 | 9504 | 3·9 |
| The Netherlands | Male | Colorectal | 0 | 0·0 | 2709 | 98·8 | 0 | 0·0 | 38752 | 98·5 | 467 | 1·1 |
| The Netherlands | Male | Stomach | 0 | 0·0 | 700 | 99·3 | 0 | 0·0 | 5100 | 98·5 | 31 | 0·5 |
| The Netherlands | Male | Liver | 0 | 0·0 | 293 | 52·4 | 0 | 0·0 | 1437 | 61·5 | 7 | 0·2 |
| The Netherlands | Male | Lung | 0 | 0·0 | 2945 | 89·4 | 0 | 0·0 | 28721 | 85·4 | 105 | 0·3 |
| The Netherlands | Male | All cancer | 0 | 0·0 | 18426 | 93·9 | 0 | 0·0 | 231264 | 94·4 | 5889 | 2·2 |
| Turin, Italy | Female | Breast | 3 | 0·5 | 608 | 97·3 | 76 | 0·8 | 9575 | 98·6 | < 5 | < 0·1 |
| Turin, Italy | Female | Colorectal | 1 | 0·5 | 178 | 88·6 | 44 | 1·2 | 3421 | 89·6 | < 5 | < 0·2 |
| Turin, Italy | Female | Cervical | 0 | 0·0 | 109 | 99·1 | 2 | 0·6 | 347 | 96·9 | < 5 | < 1·1 |
| Turin, Italy | Female | Stomach | 1 | 2·1 | 45 | 93·8 | 11 | 1·4 | 686 | 88·2 | < 5 | < 0·7 |
| Turin, Italy | Female | Liver | 0 | 0·0 | 11 | 40·7 | 23 | 5·9 | 167 | 43·2 | < 5 | < 1·3 |
| Turin, Italy | Female | Lung | 0 | 0·0 | 98 | 86·7 | 52 | 2·4 | 1801 | 83·5 | < 5 | < 0·3 |
| Turin, Italy | Female | All cancer | 12 | 0·6 | 1853 | 91·4 | 462 | 1·5 | 27875 | 90·2 | < 5 | < 0·1 |
| Turin, Italy | Male | Colorectal | 0 | 0·0 | 152 | 89·9 | 27 | 0·6 | 4454 | 92·2 | < 5 | < 0·1 |
| Turin, Italy | Male | Stomach | 0 | 0·0 | 56 | 90·3 | 15 | 1·2 | 1123 | 91·0 | < 5 | < 0·4 |

| | | | | | | | | | | | |
|---|---|---|---|---|---|---|---|---|---|---|---|
| Turin, Italy | Male | Liver | 1 | 1·8 | 33 | 57·9 | 41 | 4·1 | 443 | 44·5 | < 5 | < 0·5 |
| Turin, Italy | Male | Lung | 1 | 0·5 | 162 | 80·2 | 119 | 2·2 | 4418 | 81·3 | < 5 | < 0·1 |
| Turin, Italy | Male | All cancer | 12 | 0·8 | 1251 | 85·0 | 468 | 1·2 | 33197 | 87·4 | < 5 | < 0·1 |
| Murcia, Spain | Female | Breast | 2 | 0·6 | 307 | 99·4 | 14 | 0·4 | 3788 | 98·8 | < 5 | < 0·2 |
| Murcia, Spain | Female | Colorectal | 2 | 1·9 | 97 | 94·2 | 15 | 0·9 | 1552 | 94·5 | < 5 | < 0·3 |
| Murcia, Spain | Female | Cervical | 0 | 0·0 | 78 | 97·5 | 2 | 0·7 | 264 | 98·5 | < 5 | < 1·5 |
| Murcia, Spain | Female | Stomach | 0 | 0·0 | 23 | 95·8 | 2 | 0·5 | 341 | 92·7 | < 5 | < 1·3 |
| Murcia, Spain | Female | Liver | 0 | 0·0 | 5 | 83·3 | 0 | 0·0 | 52 | 71·2 | < 5 | < 6·4 |
| Murcia, Spain | Female | Lung | 3 | 3·5 | 77 | 89·5 | 3 | 0·5 | 545 | 91·1 | < 5 | < 0·8 |
| Murcia, Spain | Female | All cancer | 11 | 0·9 | 1128 | 94·9 | 93 | 0·7 | 12672 | 92·2 | < 5 | < 0·1 |
| Murcia, Spain | Male | Colorectal | 1 | 0·7 | 135 | 97·8 | 10 | 0·4 | 2673 | 97·1 | < 5 | < 0·2 |
| Murcia, Spain | Male | Stomach | 0 | 0·0 | 52 | 98·1 | 6 | 1·2 | 472 | 96·3 | < 5 | < 1·0 |
| Murcia, Spain | Male | Liver | 0 | 0·0 | 22 | 62·9 | 0 | 0·0 | 235 | 57·2 | < 5 | < 1·2 |
| Murcia, Spain | Male | Lung | 1 | 0·6 | 142 | 89·9 | 11 | 0·4 | 2476 | 89·2 | < 5 | < 0·2 |
| Murcia, Spain | Male | All cancer | 10 | 0·7 | 1275 | 94·4 | 99 | 0·5 | 17626 | 92·2 | < 5 | < 0·1 |
| Tarragona, Spain | Female | Breast | 0 | 0·0 | 209 | 100·0 | 9 | 0·4 | 2195 | 98·8 | 253 | 9·4 |
| Tarragona, Spain | Female | Colorectal | 0 | 0·0 | 35 | 100·0 | 18 | 1·8 | 945 | 92·9 | 257 | 19·6 |
| Tarragona, Spain | Female | Cervical | 0 | 0·0 | 31 | 100·0 | 1 | 1·2 | 80 | 98·8 | 39 | 25·8 |
| Tarragona, Spain | Female | Stomach | 0 | 0·0 | 6 | 100·0 | 4 | 3·4 | 107 | 89·9 | 54 | 30·2 |
| Tarragona, Spain | Female | Liver | | | | | 0 | 0·0 | 9 | 33·3 | 13 | n/a |
| Tarragona, Spain | Female | Lung | 0 | 0·0 | 27 | 96·4 | 4 | 1·3 | 270 | 86·8 | 150 | 30·7 |
| Tarragona, Spain | Female | All cancer | 2 | 0·4 | 472 | 97·9 | 68 | 1·1 | 5974 | 92·4 | 1457 | 17·3 |
| Tarragona, Spain | Male | Colorectal | 0 | 0·0 | 49 | 98·0 | 15 | 0·9 | 1593 | 95·2 | 417 | 19·5 |
| Tarragona, Spain | Male | Stomach | 0 | 0·0 | 12 | 92·3 | 4 | 2·0 | 192 | 93·7 | 111 | 33·7 |
| Tarragona, Spain | Male | Liver | 0 | 0·0 | 8 | 61·5 | 1 | 0·9 | 81 | 72·3 | 46 | 26·9 |
| Tarragona, Spain | Male | Lung | 0 | 0·0 | 52 | 92·9 | 7 | 0·6 | 1089 | 87·1 | 584 | 30·9 |
| Tarragona, Spain | Male | All cancer | 1 | 0·2 | 416 | 94·3 | 78 | 0·9 | 8081 | 90·9 | 2578 | 21·6 |
| Ticino, Switzerland | Female | Breast | 2 | 0·3 | 676 | 99·1 | 3 | 0·3 | 1064 | 98·4 | n/a | n/a |
| Ticino, Switzerland | Female | Colorectal | 2 | 0·8 | 244 | 98·0 | 3 | 0·8 | 353 | 95·1 | n/a | n/a |
| Ticino, Switzerland | Female | Cervical | 0 | 0·0 | 27 | 96·4 | 0 | 0·0 | 43 | 100·0 | n/a | n/a |
| Ticino, Switzerland | Female | Stomach | 0 | 0·0 | 59 | 93·7 | 3 | 4·6 | 59 | 90·8 | n/a | n/a |
| Ticino, Switzerland | Female | Liver | 0 | 0·0 | 4 | 17·4 | 0 | 0·0 | 12 | 34·3 | n/a | n/a |
| Ticino, Switzerland | Female | Lung | 1 | 0·6 | 153 | 89·0 | 4 | 1·3 | 276 | 90·8 | n/a | n/a |

| | | | | | | | | | | | | |
|---|---|---|---|---|---|---|---|---|---|---|---|---|
| Ticino, Switzerland | Female | All cancer | 19 | 0·9 | 1977 | 94·2 | 35 | 1·0 | 3184 | 94·1 | n/a | n/a |
| Ticino, Switzerland | Male | Colorectal | 1 | 0·3 | 326 | 98·2 | 1 | 0·3 | 367 | 98·4 | n/a | n/a |
| Ticino, Switzerland | Male | Stomach | 3 | 2·8 | 100 | 94·3 | 0 | 0·0 | 79 | 96·3 | n/a | n/a |
| Ticino, Switzerland | Male | Liver | 1 | 1·0 | 37 | 38·5 | 0 | 0·0 | 75 | 46·0 | n/a | n/a |
| Ticino, Switzerland | Male | Lung | 7 | 2·0 | 316 | 91·3 | 3 | 0·6 | 431 | 90·7 | n/a | n/a |
| Ticino, Switzerland | Male | All cancer | 17 | 0·7 | 2233 | 92·3 | 14 | 0·4 | 3337 | 92·5 | n/a | n/a |

DCO = death certificate only; MV = microscopically verified. N = number of cases; DCO and MV percentages use each population's own total cases as denominator.
Missing birth country = cases with no recorded country of birth; % missing = missing / (host + migrant + missing) × 100. "< 5" = fewer than five cases, withheld by the registry; n/a = not reported.
Belgium is not shown because the Belgian Cancer Registry did not share DCO and MV counts.
